# PregMedNet: Multifaceted Maternal Medication Impacts on Neonatal Complications

**DOI:** 10.1101/2025.02.13.25322242

**Authors:** Yeasul Kim, Ivana Marić, Chloe M. Kashiwagi, Lichy Han, Philip Chung, Jonathan D Reiss, Lindsay D. Butcher, Kaitlin J. Caoili, Eloïse Berson, Lei Xue, Camilo Espinosa, Tomin James, Sayane Shome, Feng Xie, Marc Ghanem, David Seong, Alan L Chang, S Momsen Reincke, Samson Mataraso, Chi-Hung Shu, Davide De Francesco, Martin Becker, Wasan M Kumar, Ron Wong, Brice Gaudilliere, Martin S Angst, Gary M Shaw, Brian T Bateman, David K Stevenson, Lawrence S Prince, Nima Aghaeepour

## Abstract

While medication use is common among pregnant women, medication safety remains insufficiently characterized because studies in pregnant women are challenging due to safety concerns. The recent digitization of healthcare databases and advances in computational methods have created new opportunities for large-scale, retrospective drug safety evaluations. Here, we present PregMedNet, a platform that characterizes multifaceted maternal medication effects on neonatal outcomes during pregnancy, covering more than 27,000 drug-disease pairs across 1,152 medications and 24 outcomes. These results encompass known and novel odds ratios (ORs), adjusted ORs, and drug-drug interactions, systematically analyzed using nationwide claims data and an advanced machine learning pipeline. Notably, one of the newly discovered associations was experimentally validated *in vivo*. This supports the reliability of PregMedNet’s findings and demonstrates the utility of claims data and machine learning for perinatal medication safety studies. Additionally, potential biological mechanisms underlying the associations were explored using a graph learning method, providing candidate pathways for future mechanistic investigations. We expect that PregMedNet will contribute to advancing maternal medication safety and improving neonatal outcomes by providing extensive, multifaceted drug safety information on this previously underrepresented population.

## Introduction

More than 80% of women use at least one medication during pregnancy, and the use of prescription medication has increased by over 60% over the last three decades.^1–3^ However, medication safety is still insufficiently characterized in pregnant women, and pregnant women and their healthcare teams frequently encounter limited or contradictory guidance on medication safety.^4^ More than one-third of newly approved drugs by the United States (US) Food and Drug Administration (FDA) during the last decade are out of compliance with the Pregnancy and Lactation Labeling Rule (PLLR), and less than 20% have adequate drug safety data on pregnancy and lactation based on human studies.^5^

One of the main reasons for the lack of medication safety evidence in pregnant women is the recognition of pregnant women as a vulnerable population and regulatory guidance has prioritized decreasing harm. Premarketing clinical trials for drug safety generally poses greater than minimal risk to pregnant women, which conflicts with FDA’s and institutional review board’s priority to provide special protections for pregnant women.^6–8^ Because of this, pregnant women have been considered “therapeutic orphans” due to the lack of adequate drug safety and efficacy data.^9–12^ The current drug safety data of pregnancy have been collected mainly through post-marketing surveillance,^13^ especially through spontaneous reporting systems (SRSs).^14^ Although SRSs are regarded as an effective tool for drug safety surveillance, they are potentially biased by under-reporting, may not be able to assess low-level risks, and are challenged to definitively quantify risks and side effects.^13–16^

Computational and data-driven approaches to assess drug safety have recently become viable fueled by rapid digitization of patient data resulting in large healthcare databases, and the development of machine learning.^10^ Large administrative claims databases are increasingly being used to assess drug safety because they contain extensive samples of real-world patient information. Unlike electronic health records (EHR) which are typically confined to a single hospital system, claims databases include patients across institutions and geographic regions, thereby providing large enough sample sizes, rich information on potential confounders, and analytic cost-effectiveness. This makes them an invaluable tool for analysis of medication effects, particularly in underrepresented populations such as pregnant women. Previous studies have demonstrated that claims databases can be successfully utilized to evaluate drug safety in pregnant women, by developing algorithms to establish a pregnant cohort and applying statistical analysis.^17–20^ However, these studies have typically concentrated on the effects of one or a few maternal medications on pregnancy outcomes.

We introduce PregMedNet, a platform dedicated to advancing maternal medication safety during pregnancy, with particular focus on neonatal complications. The impacts of multiple maternal medications used during pregnancy on neonatal outcomes were assessed based on the Merative™ Marketscan® Commercial Database,^21^ a comprehensive collection of claims data across the U.S. covering more than 250 million patients. Utilizing retrospective, large-scale mother-baby dyads, we systematically analyzed the multifaceted maternal medication impacts during pregnancy on neonatal complications with high statistical power.

First, we comprehensively analyzed 27,648 drug-disease pairs between 1,152 maternal medications and 24 neonatal complications, encompassing all medications available in the database. To minimize bias, we implemented a machine–learning–based confounding adjustment pipeline, which automatically selects and adjusts for confounding factors without requiring manual selection for each medication-complication pair. Additionally, we analyzed drug-drug interactions (DDIs), identifying additive effects of maternal drug combinations on neonatal complications. In total, 1,446 odds ratios (ORs), 261 adjusted ORs, and 5 DDIs were statistically significant, encompassing both known and novel maternal medication effects.

Notably, we experimentally validated one of the newly identified associations—between maternal ondansetron use and neonatal bronchopulmonary dysplasia (BPD)—*in vivo*. This validation supports the reliability of PregMedNet findings and highlights the potential of retrospective claims data and machine learning for maternal medication safety research.

Finally, we employed a knowledge graph and a graph learning method to uncover potential biological mechanisms underlying newly discovered associations. A case study of ondansetron and BPD demonstrated how this approach generates testable hypotheses, aiming to expedite mechanism of action (MoA) discovery by narrowing down specific molecular targets for further investigation.

Our comprehensive findings on maternal medication impacts and underlying mechanisms can be explored through an interactive website: https://pregmednet.stanford.edu/.

## Results

### Mother-baby cohort: encompassing a wide-range of variables

A total of 1,190,257 mother-baby dyads were obtained between 2007 and 2021 from the Merative™ Marketscan® Commercial Database^21^ (Supplementary Figures 1 & 2). The average maternal age was 31.18 ± 4.75 years, and the average gestational age was 38.58 ± 1.71 weeks with a preterm birth rate of 8.98% (Supplementary Table 1-1).

The discovery cohort included 817,402 dyads, used as the primary group for initial analysis and hypothesis generation. The validation cohort included 372,855 dyads, used to validate the generated hypotheses. Each cohort contained information about pregnant mothers in the 90 days prior to delivery, including 68 maternal comorbidities, preterm birth, gestational age at birth, maternal age at birth, the region and geographic location of the healthcare facilities, and all maternal medications taken during the 90 days prior to delivery. Additionally, the cohort included information about the corresponding babies during the first 90 days after birth, covering both sex and incidence of 24 neonatal complications (Figure 1.c), with prevalence presented in Supplementary Table 2. All maternal medications taken were extracted and harmonized to the medications’ compound-level identity along with route of administration information (Supplementary Figure 3). There were a total of 1,152 maternal medications in the discovery cohort and 980 in the validation cohort.

**Figure 1.**
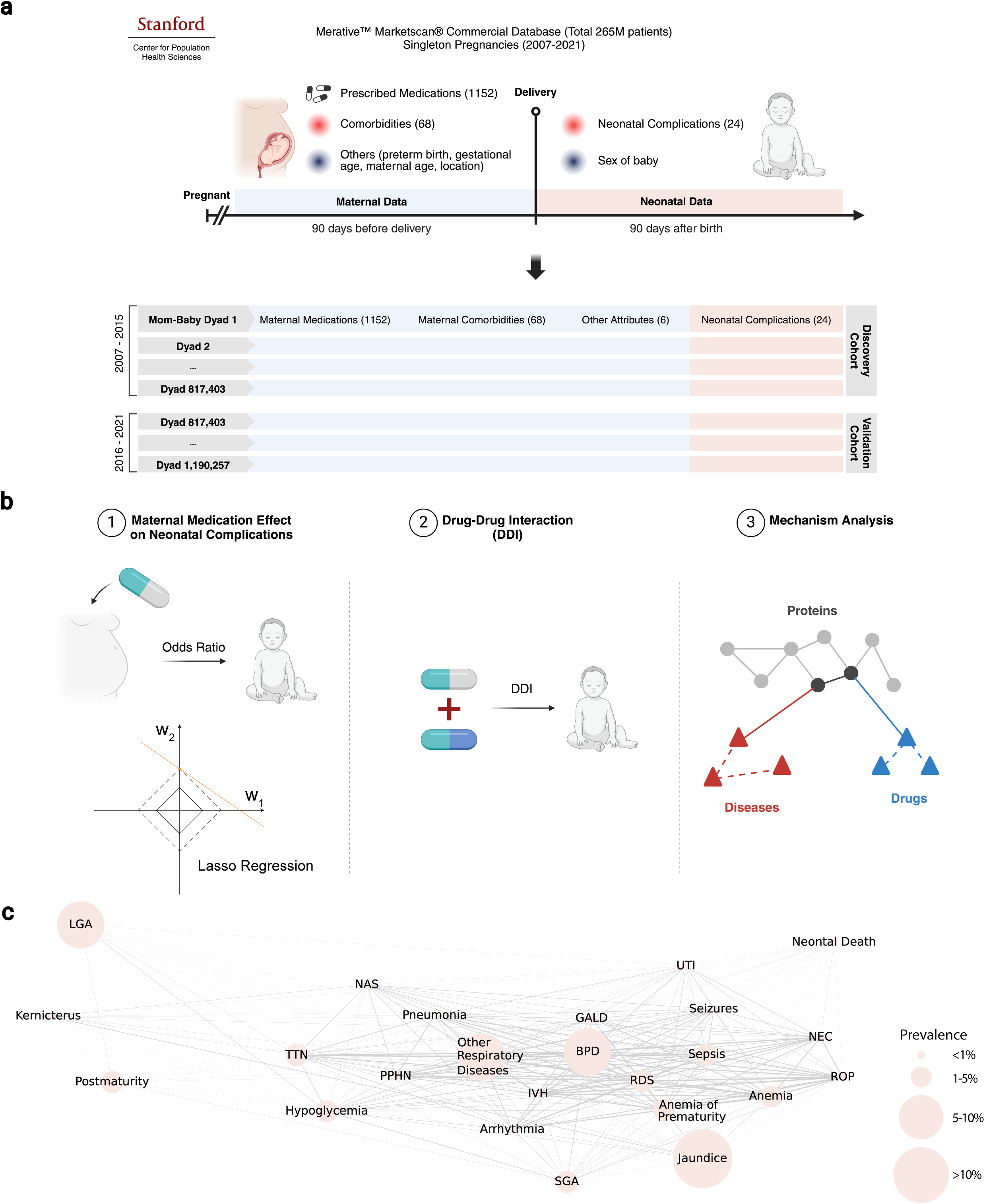
Study Overview. (a) A total of 1,190,257 mother-newborn dyads from 2007 to 2021 are extracted from the Merative™ Marketscan® Commercial Database, a set of de-identified claims data accessed through the Stanford Center for Population Health Sciences. Maternal records from the last 90 days before pregnancy are examined to extract 68 maternal comorbidities, 6 additional attributes, and all prescribed medications. Similarly, newborn records from the first 90 days after birth are analyzed to identify 24 neonatal complications. The final dataset includes 1,152 maternal medications, 68 maternal comorbidities, 24 neonatal complications, and 6 additional attributes: maternal age, gestational age at delivery, preterm birth, baby’s sex, and the region and geographic location of the healthcare facility. The dyads are divided into a discovery cohort (2007-2015) and a validation cohort (2016-2021) based on the year of birth. (b) Three experiments are conducted based on the database. First, the associations between maternal medications and neonatal complications are calculated using odds ratios based on logistic regression. Notably, confounders are selected and adjusted for each medication-disease pair using the LassoNoExp algorithm, which penalizes and selects confounding variables using Lasso regression while preserving maternal medication exposure information. Second, additive effects of concomitant maternal medications on neonatal complications are calculated through drug-drug interaction analysis. Lastly, potential underlying mechanisms of newly discovered drug-disease associations are discovered using a knowledge graph and a graph learning method. (c) A tetrachoric correlation plot of the 24 neonatal outcomes shows the relationships between the neonatal complications. The size of each node is correlated with disease prevalence, and the nodes are connected if the diseases are positively correlated. The edge thickness is proportional to the strength of the correlation.thickness is proportional to the strength of the correlation.

### Multifaceted insights into perinatal medication safeties

PregMedNet offers comprehensive insights into the impacts of maternal medications on neonatal complications, examining (a) effects of single medications, (b) impacts of concomitant medications on neonatal outcomes, and (c) potential underlying mechanisms behind significant associations.

### Individual maternal medication impacts on newborn outcomes

The associations between maternal medications and neonatal complications were investigated by estimating both unadjusted and adjusted odds ratios (ORs and aORs, respectively). Out of 27,648 medication-complication pairs, 1,446 ORs (5.23%) and 261 aORs (0.94%) were statistically significant, encompassing both already known and novel maternal medication effects on neonatal complications.

#### Network graphs visualize intricate patterns of associations

PregMedNet illustrates these associations using network graphs, with ORs presented in Figure 2 and aORs in Supplementary Figure 4. Medications and diseases are represented as nodes clustered by groups, with edges color-coded to indicate increased risk (gray, OR or aOR>1) or reduced risk (blue, OR or aOR<1) associations. The network graphs highlight intricate patterns between medications and diseases.

**Figure 2.**
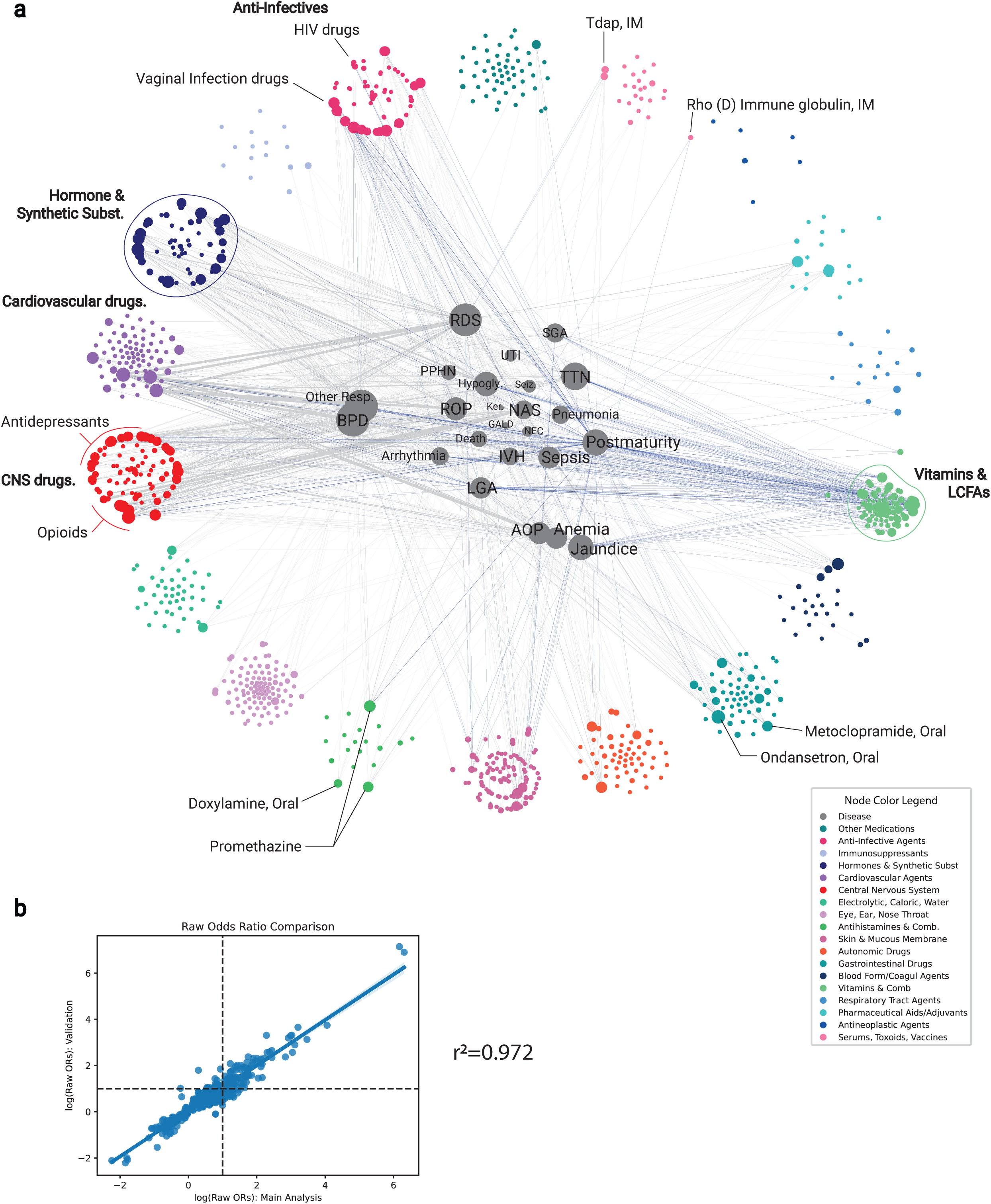
A Network of the Associations Between Maternal Medications and Neonatal Complications, and Their Validation. (a) Odds Ratio (OR) results are displayed as a network graph, where nodes represent maternal medications or neonatal complications, and edges indicate statistically significant ORs. Disease nodes are centrally placed, while maternal medication nodes are peripheral. Medications are grouped and color-coded within each cluster. The graph shows significant ORs as edges; gray for ORs above 1 (positive correlations), and blue for ORs below 1 (negative correlations). There are 24 disease nodes, 1,152 medication nodes, and 1,446 edges—1,153 gray and 293 blue. Edge thickness correlates with the strength of the association (inverse of p-values), and node size with the number of connecting edges. Abbreviations in disease nodes: BPD: Bronchopulmonary Dysplasia, Other Resp.: Other Respiratory Diseases, RDS: Respiratory Distress Syndrome, SGA: Small for Gestational Age, PPHN: Persistent Pulmonary Hypertension, Hypogly.: Hypoglycemia, UTI: Urinary Tract Infection, Seiz.: Seizures, TTN: Transient Tachypnea, ROP: Retinopathy of Prematurity, Ker.: Kernicterus, GALD: Gestational Alloimmune Liver Disease, NAS: Neonatal Abstinence Syndrome, NEC: Necrotizing Enterocolitis, IVH: Intraventricular Hemorrhage, Death: Neonatal Death, LGA: Large for Gestational Age, AOP: Anemia of Prematurity. Additional conditions in disease nodes not abbreviated: Pneumonia, Postmaturity, Sepsis, Arrhythmia, Anemia, and Jaundice. (b) ORs from the discovery cohort were validated in the validation cohort. ORs for both cohorts were matched by maternal medications and neonatal complications and plotted on a scatterplot. This figure compares the log of significant ORs from the discovery (x-axis) to the validation cohort (y-axis). Of the ORs, 772 matched between the cohorts—1,446 significant in the discovery cohort and 1,097 in the validation cohort. Pearson correlation yielded an R² of 0.972 with a p-value of 0.0.

In the OR network graph (Figure 2), 1,153 edges (79.7%) are gray, while 293 edges (20.3%) are blue, out of a total of 1,446 edges. Focusing on diseases, neonatal respiratory diseases, particularly Respiratory Distress Syndrome (RDS) (140 gray edges) and BPD (135 gray edges), exhibit the most increased risk associations with maternal medications. In contrast, Postmaturity (87 blue edges) and Large for Gestational Age (49 blue edges) show the most reduced risk associations. Focusing on maternal medication groups, the Central Nervous System (CNS) group has the highest number of increased risk associations (300 gray edges), followed by the Hormone and Synthetic Substances group (212 gray edges). Conversely, the Vitamins & Combinations group shows the highest number of reduced risk associations (151 blue edges), followed by the Anti-Infective Agents group (40 blue edges). Supplementary Figure 5 provides detailed information on the number of significant ORs per neonatal complication and maternal medication group.

In contrast, the aOR network graph (Supplementary Figure 4) reveals a shift toward decreased risk associations, with 117 edges (44.8%) gray and 144 edges (55.2%) blue out of 261 total edges. Focusing on diseases (Figure 3.a.), patterns with aORs align with ORs: neonatal respiratory complications show the highest increased risk associations, led by BPD (24 gray edges), followed by Other Respiratory (21 gray edges) and RDS (19 gray edges), while Postmaturity (30 blue edges) and large-for-gestational age (LGA) (23 blue edges) have the most decreased risk associations. Similarly, focusing on maternal medication groups (Figure 3.b.), CNS medications show the highest increased risk associations (50 gray edges), followed by Cardiovascular Agents and Hormone and Synthetic Substances (14 gray edges for each), while Vitamins & Combinations show the most decreased risk associations (102 blue edges), followed by Anti-Infective Agents (13 blue edges). Supplementary Figure 6 provides detailed counts of significant aORs per neonatal complication and maternal medication group.

**Figure 3.**
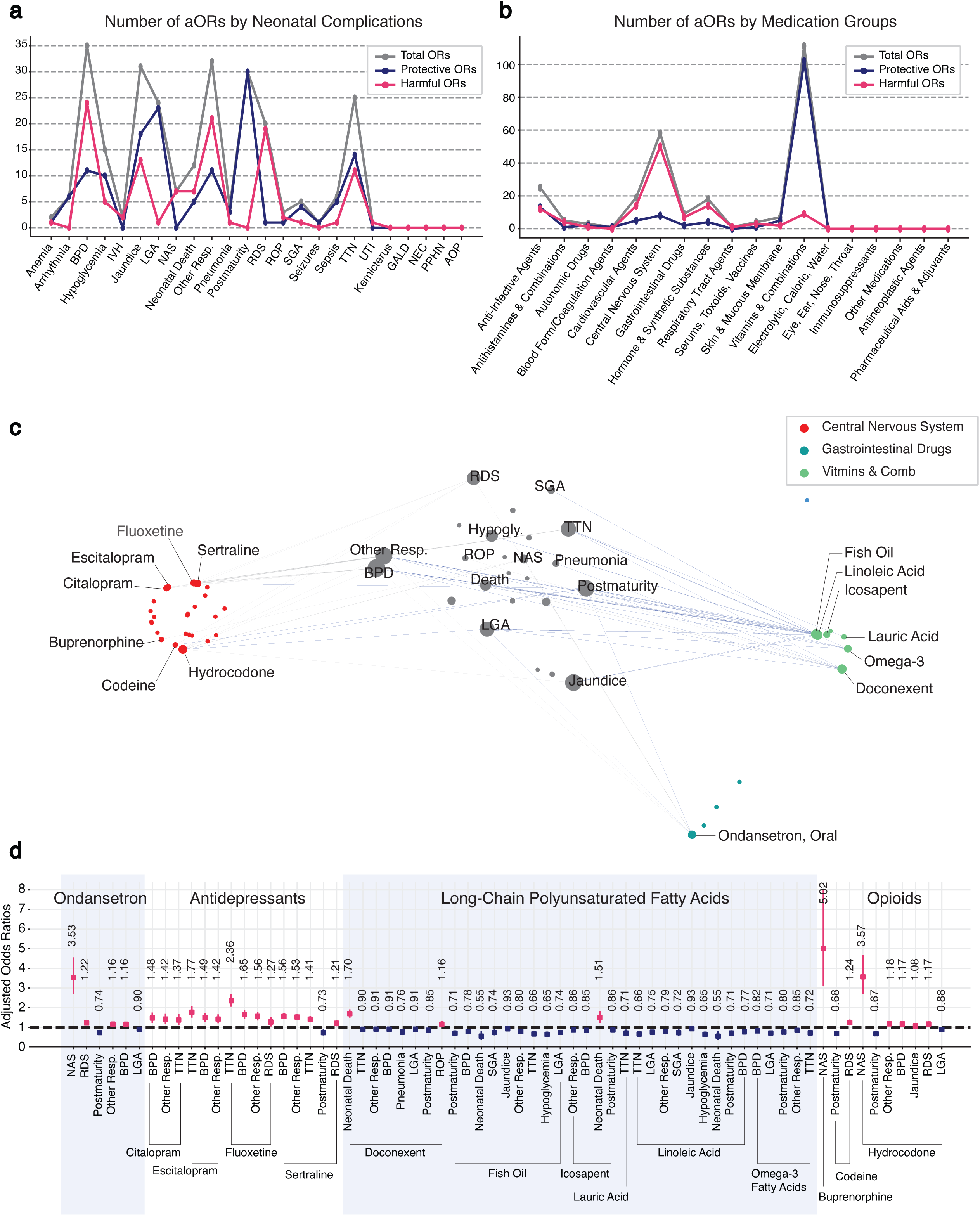
Associations Between Four Maternal Medication Groups and Neonatal Complications: Highlights from the Adjusted Odds Ratio Network. (a) The figure shows the number of aORs by neonatal complications. The gray line represents the total aORs, the navy line indicates reduced risk aORs, and the magenta indicates increased risk aORs. (b) The figure shows the number of aORs by maternal medication groups, with the line colors corresponding to those in (a). (c) This network graph highlights four maternal medication groups and their adjusted odds ratios (aOR) with neonatal complications. Nodes are consistent with those in Figure 2’s unadjusted odds ratio graph, while edges represent statistically significant adjusted odds ratios. LassoNoExp was utilized to calculate these aORs, selecting and adjusting for maternal covariates out of a possible 73 for each drug-complication pair. Edges in gray indicate aORs greater than 1 (positive correlations), and those in blue indicate aORs less than 1 (negative correlations). The edge thickness corresponds to the association strength, inversely proportional to the p-values. Highlighted medication groups include Ondansetron, Antidepressants, Long-Chain Polyunsaturated Fatty Acids (LC-PUFAs), and Opioids. The full network can be examined in Supplementary Figure 6. (d) The subfigure details the values of statistically significant adjusted odds ratios and their confidence intervals for the four medication groups. Magenta dots and lines depict aORs higher than 1, signifying positive correlations, whereas navy blue dots and lines indicate aORs lower than 1, representing negative correlations.

#### Detailed information on associations: highlighted examples of ORs

PregMedNet provides detailed ORs and aORs for all medication-disease pairs through an interactive website, presenting both significant and non-significant results. Supplementary Figure 7 highlights key examples among 1,446 significant ORs.

Notably, within the CNS category, antidepressants and opioids are associated with elevated ORs for several neonatal complications, such as neonatal respiratory disorders and neonatal abstinence syndrome (NAS) (Supplementary Figures 7.a, 7.b). Similarly, antiemetics - commonly used during pregnancy^22^ - are associated with elevated ORs for multiple neonatal complications (Supplementary Figure 7.c). In contrast, long-chain polyunsaturated fatty acids (LC-PUFAs) in the Vitamins & Combinations category are highlighted here, showing substantial reduced risk associations for multiple neonatal complications including neonatal respiratory disorders (Supplementary Figure 7.d). Medications in the Anti-Infective Agents category also exhibit reduced risk associations (Supplementary Figure 7.e), with oral valacyclovir linked to reduced risks across multiple neonatal complications, consistent with prior studies.^17^

#### aORs derived through machine-learning-based confounder adjustment

While ORs provide valuable insights, they are unadjusted for potential confounders, which could introduce bias or spurious associations. In PregMedNet, a machine-learning-based method that selects and adjusts for confounding variables across multiple disease-medication pairs was employed, systematically estimating aORs. In the following section, we focus on four medication groups: ondansetron, antidepressants, LC-PUFAs, and opioids.

##### Ondansetron linked to increased neonatal respiratory risks

In PregMedNet, ondansetron is categorized into three types based on the route of administration: oral, oromucosal, and injection. Among these, orally administered ondansetron exhibits six statistically significant aORs, including three elevated risk associations for neonatal respiratory complications: RDS (aOR 1.22, 95% CI 1.13 – 1.32), BPD (aOR 1.16, 95% CI 1.09 – 1.23), and other respiratory diseases (aOR 1.16, 95% CI 1.09 – 1.23). Comprehensive estimates are presented in Table 1, Figure 3.c-d.

**Table 1.**
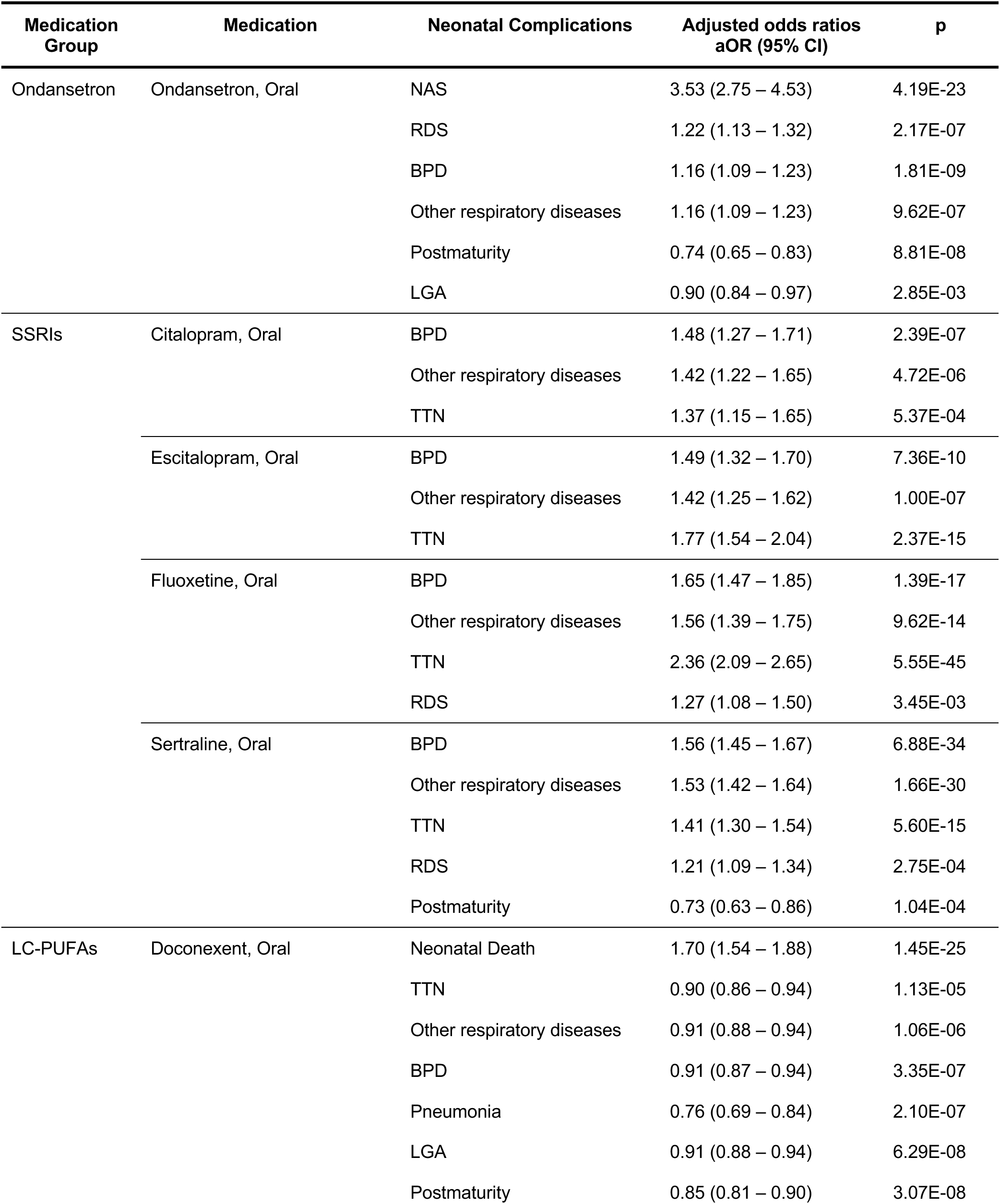
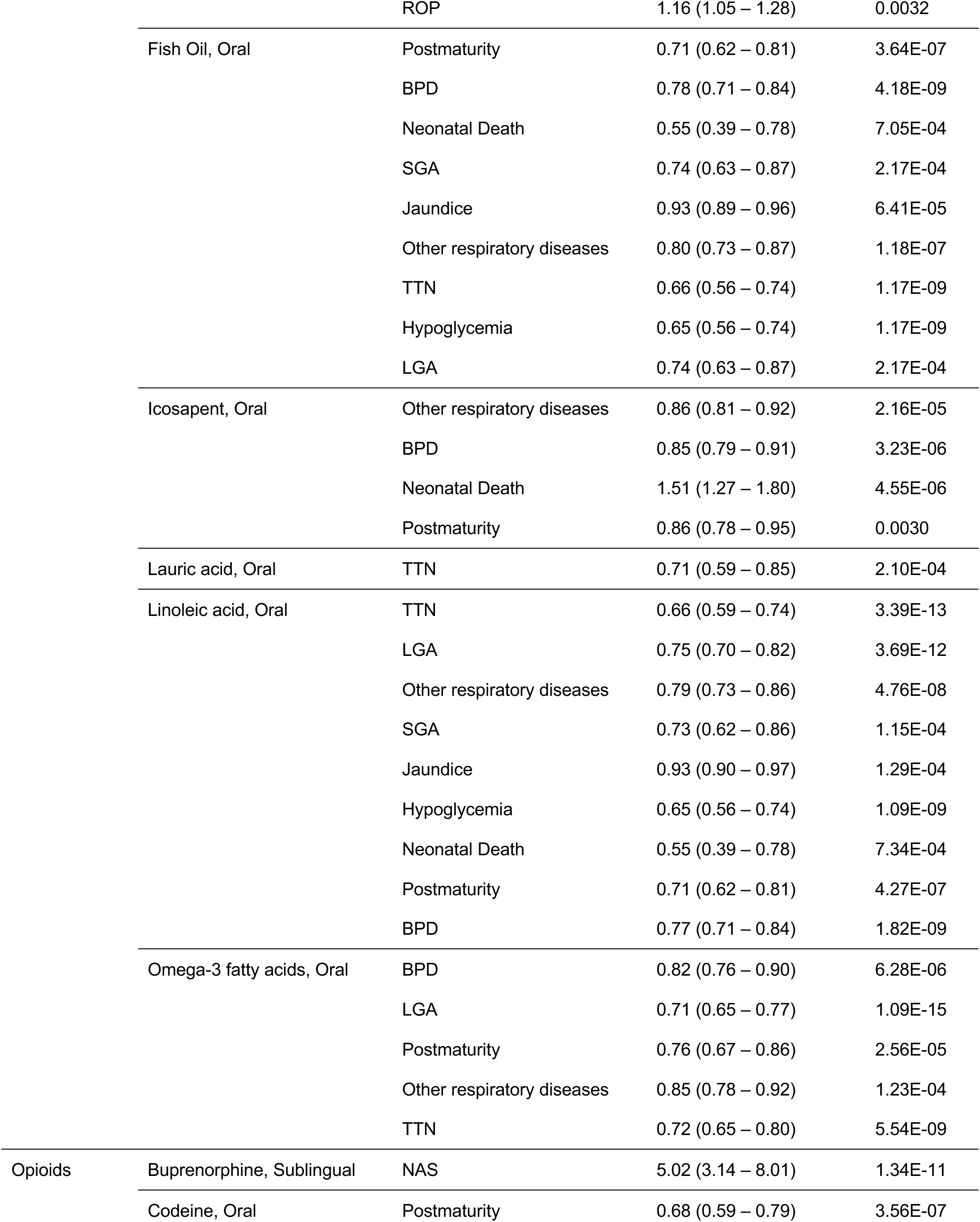
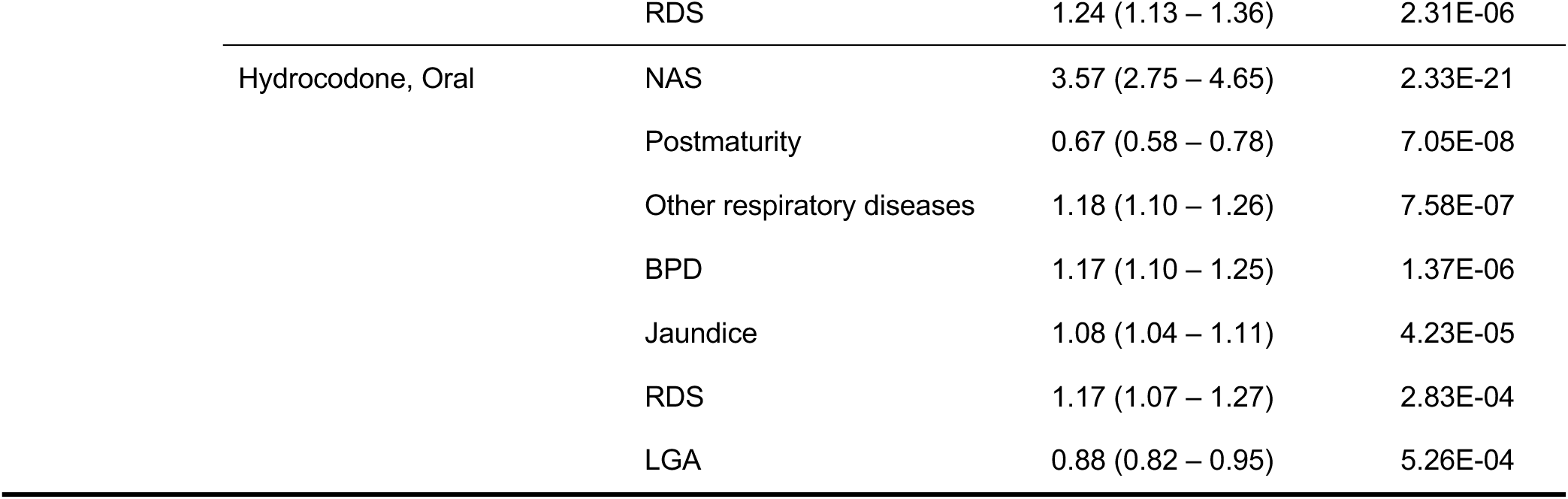
Adjusted Odds Rations (aORs) for Four Medication Groups. This table presents selected aORs from four medication groups highlighted in the Result section. The complete set of 261 significant associations is available in the interactive website (http://pregmednet.stanford.edu). Abbreviations: SSRIs: Selective Serotonin Reuptake Inhibitors, LC-PUFAs: long-chain polyunsaturated fatty acids, NAS: Neonatal Abstinence Syndrome, RDS: Respiratory Distress Syndrome, BPD: Bronchopulmonary Dysplasia, Other respiratory diseases: Other respiratory diseases, Postmaturity: Postmaturity, LGA: Large for Gestational Age, Neonatal Death: Neonatal Death, Pneumonia: Pneumonia, ROP: Retinopathy of Prematurity, SGA: Small for Gestational Age, Jaundice: Jaundice, Hypoglycemia: Hypoglycemia

Ondansetron is a medication used to treat nausea and vomiting during pregnancy, with a prevalence exceeding 25% in the US.^23^ The American College of Obstetricians and Gynecologists (ACOG) recommends cautious use of ondansetron during pregnancy due to insufficient safety data, as current studies are limited by small sample sizes and potential recall bias ^24^. The European Medicines Agency’s Pharmacovigilance Risk Assessment Committee (EMA PRAC) advises against the use of ondansetron during the first trimester of pregnancy due to recent studies that have found some associations with congenital malformations.^25, 26^ Our study, based on a large-scale retrospective database, addresses some of these limitations and found that maternal use of ondansetron during the last 90 days before delivery is associated with neonatal complications, particularly those related to the neonatal respiratory system.

##### Neonatal respiratory risks reconfirmed in perinatal SSRI use

Four SSRIs — citalopram, escitalopram, fluoxetine, and sertraline, all administered orally — show increased associations with neonatal respiratory complications. For example, citalopram presented increased associations with BPD (aOR 1.48, 95% CI 1.27 – 1.71), other respiratory disorders (aOR 1.42, 95% CI 1.22 – 1.65), and transient tachypnea of newborn [TTN] (aOR 1.37, 95% CI 1.15 – 1.65). Similar associations were observed for the other SSRIs, as detailed in Table 1 and Figure 3.c-d, and these findings are consistent with previous research results.^27, 28^

Approximately 10% of pregnant women experience depression and 2-3% take antidepressants, yet their use during pregnancy is controversial due to potential adverse effects on neonates.^29^ Selective serotonin reuptake inhibitors (SSRIs) are the most commonly used antidepressants during pregnancy, because they have a more favorable safety profile than other antidepressant drug classes. However, findings associate them with neonatal complications,^29–31^ including an increased risk of respiratory complications in neonates.^27, 28^ PregMedNet reconfirms the increased risk of neonatal respiratory complications in perinatal SSRI-exposed babies based on aORs. Specifically, PregMedNet is able to identify associations at the level of individual medications rather than SSRI medications as a whole, thus providing more granular safety information for each medication.

##### LC-PUFAs linked to reduced risks across neonatal outcomes

LC-PUFAs, categorized under Vitamins & Combinations, exhibited the most significant reduced risk associations with neonatal complications, such as BPD, other respiratory diseases, and TTN, with six out of seven LC-PUFAs showing reduced risk effects.

Fish oil and linoleic acid are particularly noteworthy for their reduced risk associations, each displaying nine reduced risk aORs related to various neonatal complications, including disorders of the respiratory system (BPD, other respiratory diseases, TTN), gestational age or size-related disorders (both LGA and SGA, postmaturity), hypoglycemia, jaundice, and neonatal death. For instance, linoleic acid’s reduced risk associations include BPD (aOR 0.77, 95% CI 0.71 – 0.84), LGA (aOR 0.75, 95% CI 0.70 – 0.82), hypoglycemia (aOR 0.65, 95% CI 0.56 – 0.74), and neonatal death (aOR 0.55, 95% CI 0.39 – 0.78). Fish oil showed a comparable pattern, with nine reductions across the same domain; the remaining results are provided in Table 1 and Figure 3.c-d.

Previous studies have shown the associations between maternal LC-PUFAs levels during pregnancy and maternal health, including preeclampsia and postpartum depression,^32, 33^ as well as their association with children’s health, including neurodevelopmental outcomes.^34^ Furthermore, recent research suggests that LC-PUFAs may also decrease the incidence of asthma and wheezing in infants, although this evidence remains limited.^35^ PregMedNet highlights the perinatal benefits of LC-PUFAs in addressing neonatal respiratory disorders, reinforcing previous findings while uncovering previously unknown associations, such as hypoglycemia and neonatal death.

##### Opioid use associated with multiple neonatal complications

As many as 6.6% of pregnant women use prescription opioids and there is concern these opioids can cause neonatal complications.^36^ PregMedNet found associations between opioids and increased risks of serious neonatal complications. For example, maternal oral hydrocodone exhibited seven increased risk associations, including NAS (aOR 3.57, 95% CI 2.75 – 4.65) and BPD (aOR 1.17, 95% CI 1.10–1.25), with further outcomes detailed in Table 1 and Figure 3.c-d.

The medications highlighted in this section identified both previously reported and novel associations as ORs and aORs, demonstrating PregMedNet’s reliability and efficiency in assessing maternal medications’ impact on neonatal complications. While this section highlights selected medication groups, comprehensive results, including all 1,446 ORs and 261 aORs, are available on our interactive website (http://pregmednet.stanford.edu/).

#### Systematic Analysis of Maternal Drug-Drug Interactions

PregMedNet also captures the additive effects of concurrent use of maternal medications on neonates through a systematic analysis of Drug-Drug Interactions (DDIs) for all possible maternal medications pairs out of 1,152 medications and their effects on 24 neonatal complications. 𝑂𝑅*_1_*, 𝑂𝑅*_2_*, and 𝑂𝑅*_12_* represent the ORs of medication 1, medication 2, and the concurrent use of two medications, respectively, while 𝛽*_3_* represents the contribution of the DDI to the overall associations between the concurrent use of medications and the neonatal complications. Negative 𝛽*_3_* suggests that the interaction reduces the risk of neonatal complications, while the positive 𝛽*_3_* suggests it increases the risk. The magnitude of association is proportional to the absolute value of 𝛽*_3_*. (Figure 4)

**Figure 4.**
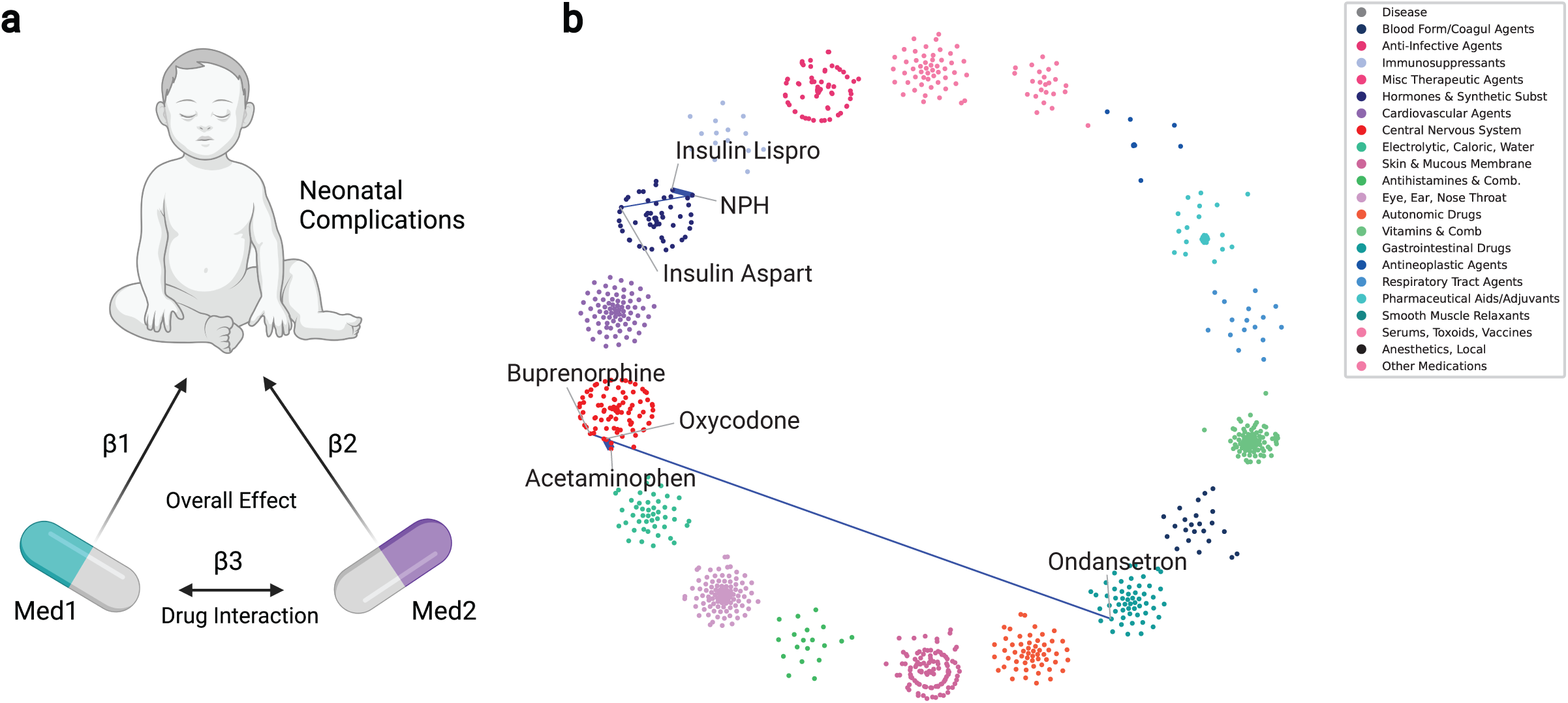
Maternal Drug-Drug Interaction Effects on Neonatal Complications. (a) Drug-Drug Interaction (DDI) effects from the concomitant use of maternal medications were calculated based on mother-baby dyads. Here, β_1_, β_2_, and β_3_ represent the individual contributions of the medications and their interaction to the overall effect on neonatal complications. Specifically, β_1_ represents the impact of medication 1, β_2_ the impact of medication 2, and β_3_ the interaction contribution between medications 1 and 2 to the collective effect on neonatal complications. (b) A network graph illustrates the DDIs as edges connecting the medication nodes. Notably, five DDIs significant in both discovery and validation cohorts are illustrated. All five edges are colored blue, indicating that the DDIs reduce neonatal complications, thereby offering additional protective benefits through the combination of maternal medications.

A total of 72 significant DDIs are identified in the discovery cohort and 15 in the validation cohort with 5 DDIs demonstrating significant 𝛽*_3_* and 𝑂𝑅*_12_* values in both the discovery and validation cohorts. All five medication pairs exhibit negative 𝛽*_3_* values, indicating that the total effect of use of two medications is less than the additive effects of two medications (Table 2).

**Table 2.** Maternal Drug-Drug Interaction Effects on Neonatal Complications. This table details the five significant Drug-Drug Interactions (DDI). The term 𝛽_3_ represents the contribution of the DDI to the overall effect of medication pairs on a neonatal complication. 𝑂𝑅*_1_*, 𝑂𝑅*_2_*, and 𝑂𝑅*_12_* represent the effect of medication 1, medication 2, and the concurrent use of two medications on a neonatal complication, respectively. A negative 𝛽_3_ indicates the reduced risk effects of the DDI on neonatal complications.

| Disease | Medication 1 | Medication 2 | $\beta_3$ | $OR_1$ | $OR_2$ | $OR_{12}$ |
| --- | --- | --- | --- | --- | --- | --- |
| NAS | Acetaminophen, Oral | Oxycodone, Oral | -3.50 | 4.89 | 99.97 | 14.77 |
| Hypoglycemia | NPH, subcutaneous | Insulin Lispro, Injection | -1.84 | 2.19 | 5.92 | 2.07 |
| Hypoglycemia | NPH, subcutaneous | Insulin Aspart, Injection | -1.52 | 2.14 | 5.42 | 2.55 |
| NAS | Buprenorphine, sublingual | Ondansetron, Oral | -1.89 | 607.34 | 5.17 | 474.43 |
| RDS | NPH, subcutaneous | Insulin Aspart, Injection | -1.02 | 1.44 | 2.25 | 1.17 |

One of the examples is the concurrent use of oral oxycodone and oral acetaminophen. The DDI effect for this combination on NAS was 𝛽_3_=-3.50, with 𝑂𝑅_12_=14.77, which was lower than the risk obtained by adding the effects of each medication alone (oxycodone: 𝑂𝑅_1_=99.97, acetaminophen: 𝑂𝑅_2_ =4.89). This attenuation indicates that concurrent use may modify neonatal risk profiles through DDI. Prior research has shown that combining oxycodone with other analgesics–NSAIDs– improves pain management over oxycodone alone.^37^ In a similar manner, our findings highlight the potential implications of oxycodone in combination with other analgesics in the perinatal population.

Another medication pair to highlight is the neonatal benefits of concurrent use of maternal insulins. The DDI effect of combined use of maternal Insulin Human Isophane (NPH, subcutaneous) and Insulin lispro (injection) for neonatal hypoglycemia is 𝛽*_3_* = -1.84, and the 𝑂𝑅*_12_* = 2.03. Notably, the 𝑂𝑅*_12_* is lower than the OR for each single medication (NPH: 2.19, Insulin lispro: 5.97), indicating that the concurrent use is associated with an overall lower risk of neonatal hypoglycemia. Similarly, the DDI effect of the combination of NPH (subcutaneous) and Insulin aspart (injection) on hypoglycemia is 𝛽*_3_* =- 1.52 with 𝑂𝑅*_12_* = 2.55, which is lower than the odds observed for Insulin Aspart alone (NPH: 𝑂𝑅_1_ =2.14, Insulin Aspart: 𝑂𝑅_2_ =5.42). These findings suggest that basal-bolus insulin regimens for the management of gestational diabetes (GDM),^38^ result in superior maternal glucose control compared to using a single insulin regimen. Improved glucose regulation minimizes in-utero fetal hyperinsulinemia, a common complication in GDM, ultimately reducing the risk of neonatal hypoglycemia at birth.

Lastly, there are two additional medication pairs which show significant 𝛽*_3_* in both the discovery cohort and the validation cohort: Buprenorphine (sublingual) and ondansetron (oral) for NAS (𝛽*_3_* =- 1.89), and NPH (subcutaneous) and Insulin aspart (injection) for RDS (𝛽*_3_* =-1.02).

The DDI analysis conducted through PregMedNet shows that certain well-known medication pairs provide additional benefits for neonatal complications when used concurrently in mothers. Although we have spotlighted five medication pairs with overlapping significance in both the discovery and validation cohorts, it is important to note that DDIs identified exclusively in one cohort still hold value. Further details on the additional DDIs are available on our interactive website.

#### Validation of Single and Concomitant Medication Effects

The ORs and aORs, along with DDIs results from the discovery cohorts, were validated in the independent validation cohort using the same analysis. Scatter plots and Pearson correlation coefficients (Figure 2.b and Supplementary Figure 8) demonstrated strong consistency (𝑅*^2^* = 0.972 for ORs, 𝑅*^2^* = 0.747 for aORs, and 𝑅*^2^* = 0.920 for DDIs). These results indicate maternal medication associations derived from the discovery cohort were reproducible in the independent cohort across differing conditions, such as study year, suggesting robustness of PregMedNet findings.

#### Maternal ondansetron impairs neonatal lung development *in vivo*

To further evaluate the robustness of the newly discovered associations, we experimentally investigate the novel association, the effect of maternal ondansetron on fetal and neonatal lung development *in vivo*. Ondansetron was of particular interest due to its high usage during pregnancy and uncertain perinatal safety.^24, 25^ Our analysis showed that maternal ondansetron use was associated with increased odds of neonatal BPD (aOR = 1.16, p-value = 1.18e-6), warranting further investigation.

Timed pregnant C57BL/6 female mice were dosed daily with 5 mg/kg bodyweight ondansetron by intraperitoneal injection from embryonic day 15 (E15) until delivery. Lung histology was assessed on postnatal day 11 (P11) to test if ondansetron exposure might impact saccular and alveolar stage lung development (Figure 5). Maternal ondansetron exposure did not affect neonatal survival or weight gain (Figures 5 b,c), suggesting that secondary influences on lung development can be excluded. However, the lungs of ondansetron exposed mice had increased alveolar mean linear intercept, indicative of reduced alveolar septation and interstitial septal thinning ^39^ (Figure 5d,e). These findings support an association between maternal ondansetron exposure and subsequent neonatal lung abnormalities, consistent with the associations identified through PregMedNet.

**Figure 5:**
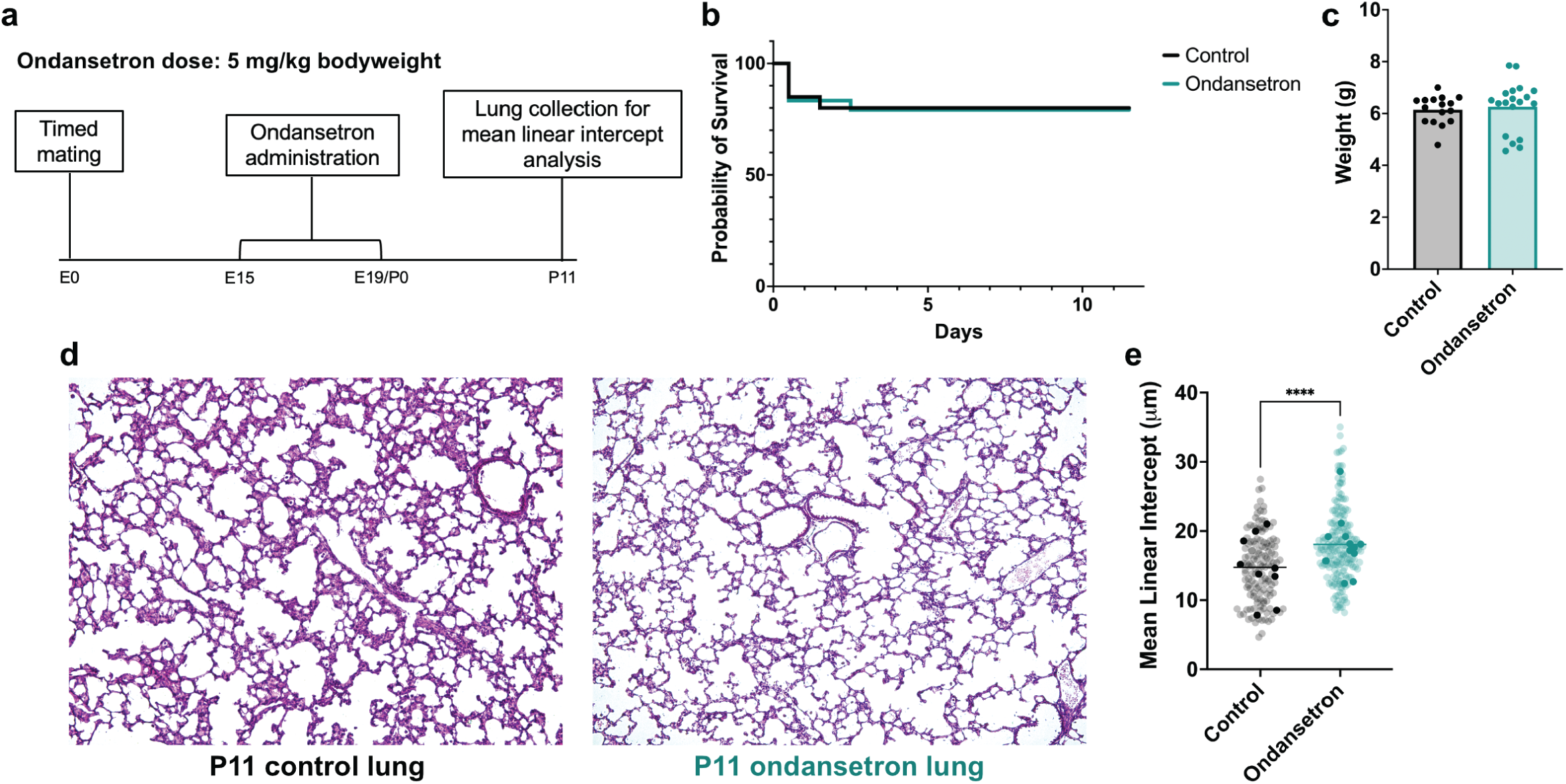
Impact of maternal ondansetron exposure and neonatal lung development in mice. (a) Experimental timeline of maternal ondansetron administration and quantification of lung development. Timed pregnant C57BL/6 female mice were injected daily with ondansetron (5 mg/kg i.p.) or an equivalent volume of sterile PBS (control) from E15 until delivery. (b) Kaplan-Meier survival analysis and (c) neonatal weight of control and ondansetron exposed mice. N = 20 mice from 4 litters in the control group; n = 24 mice from 5 litters in the ondansetron group. (d) Representative H&E images of lungs from control and ondansetron exposed mice at postnatal day 11. (e) Mean linear intercept quantification of control and ondansetron exposed mice. For each mouse, 3 lung sections were stained, with 5 random images taken from each section for a total of 135-180 images from 9-12 mice. Darker, overlayed dots represent average mean linear intercept values for each individual mouse. **** indicates P < 0.0001 as determined using an unpaired t test.

#### Knowledge Graph Reveals Potential Underlying Mechanisms

PregMedNet identifies potential underlying mechanisms of the newly discovered associations between maternal medications and neonatal complications. This was achieved by combining a knowledge graph^40^ and subsequently applying a graph learning method for the 261 significant aORs, which drew associations between 19 neonatal complications and 68 maternal medications. Of these, 15 neonatal complications and 63 maternal medications were mapped from PregMedNet to corresponding nodes in the knowledge graph, resulting in 153 disease-medication pairs for which mechanistic hypotheses are tested.

Two types of subgraphs were constructed: one containing all biological interactions and the other one with only proteins/genes between the medication and disease pair. Hypothesis testing involved extracting the shortest paths between the medication and the disease nodes and examining the involved biological nodes on these paths. This process was iterated across all 153 associations.

Notably, the mechanistic pathways underlying maternal ondansetron’s effect on neonatal BPD are highlighted here as a case study. PregMedNet generates plausible hypotheses involving interleukin-1b (IL-1b), known as a key factor in BPD pathophysiology.^41, 42^ Hypotheses based on the protein/gene-only subgraph (Figure 6) are particularly insightful, as they only include direct molecular interactions, avoiding non-specific biological entities (e.g., generic “protein bindings”), while the full biological interaction subgraphs (Supplementary Figure 9) generates a broader range of hypotheses.

**Figure 6.**
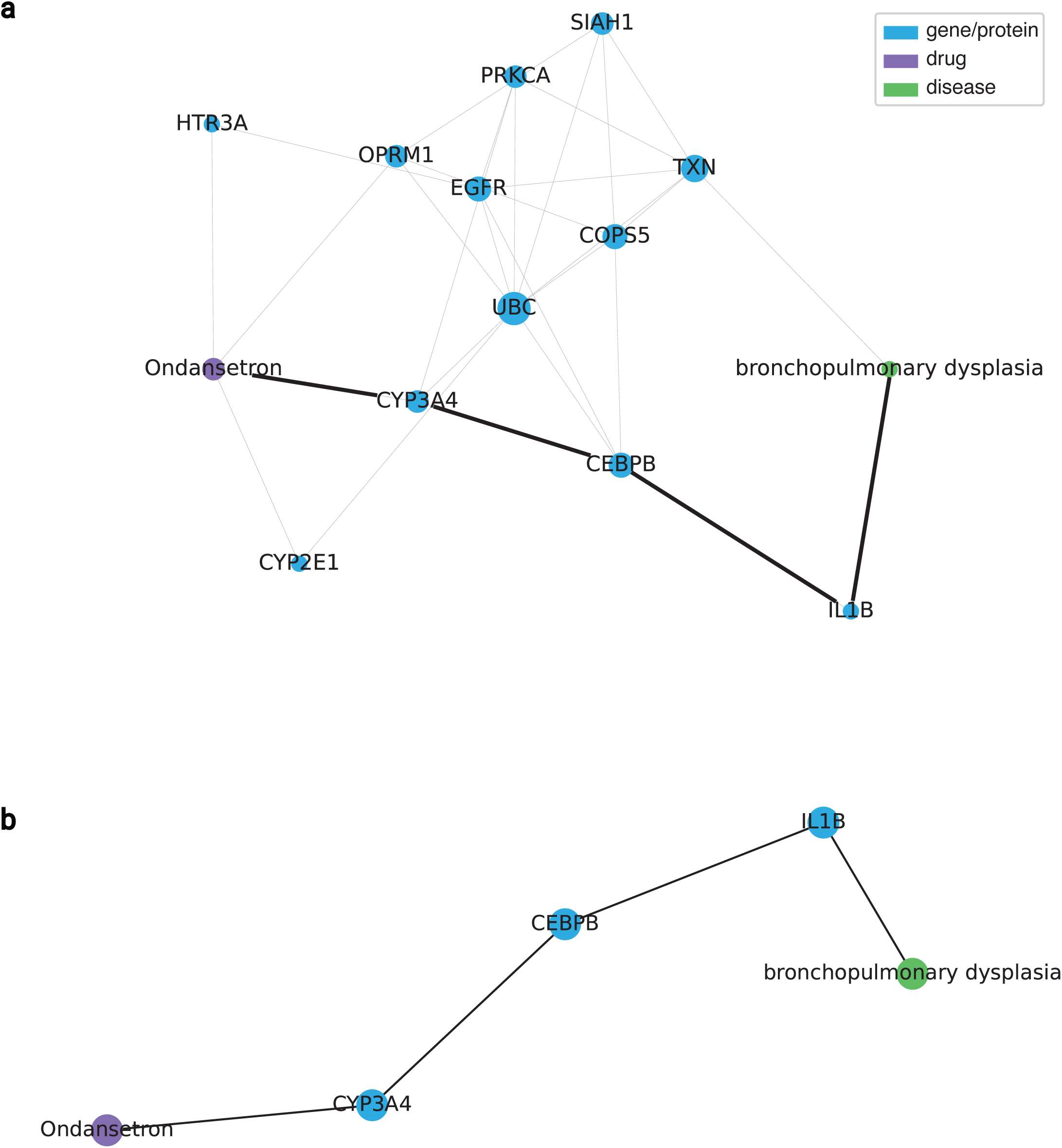
Potential Underlying Mechanisms of Ondansetron Effects on Bronchopulmonary Dysplasia. (a) This figure illustrates the potential underlying mechanisms behind the newly discovered associations between maternal medications and neonatal complications, utilizing significant adjusted odds ratios from PregMedNet and the PrimeKG Knowledge Graph, which contains scientifically validated biological interactions. Specifically, the subgraph containing three different node types—drugs, proteins/genes, and diseases—is shown here to generate more reliable hypotheses than the other type of subgraph containing additional biological node types. Displayed is the subgraph of all nodes and edges that connect ondansetron to bronchopulmonary dysplasia via the shortest paths between the two nodes. This results in eight paths with twelve intermediary nodes. Each path represents a hypothesis for a possible underlying mechanism of action. (b) A particular path of interest suggests that ondansetron affects bronchopulmonary dysplasia through the protein interactions between CYP3A4 and IL-1b. This hypothesis builds on previous research showing the relationship between ondansetron and CYP3A, and a separate relationship between bronchopulmonary dysplasia and IL-1b. While there is no prior published literature that relates CYP3A4 to IL-1b in the context of ondansetron and bronchopulmonary dysplasia, PregMedNet can generate hypotheses for novel relationships by connecting already proven, but isolated biological knowledge using graph learning methods.

PregMedNet suggests that the shortest paths between ondansetron and BPD consist of 4-hop paths, meaning it takes four edges to reach the other side of the graph (Figure 6.a.). A total of 8 shortest paths were identified, involving 12 intermediary nodes. Each path highlights potential mechanisms, visually represented by the edges between the nodes in the graph.

One of these paths suggests that ondansetron and BPD are connected via an interaction between IL-1b and Cytochrome P450 3A4 (Figure 6.b). While there are prior research studies linking IL-1b to arrested lung development in both human and experimental BPD^41, 42^, ondansetron to multiple forms of cytochrome P450 (CYP450)^43, 44^, and CYP450 to IL-1b ^45^, to the best of our knowledge, no prior research has investigated the mechanism by which maternal ondansetron might impact neonatal BPD risk. As such, PregMedNet, in this instance, generates testable hypotheses by connecting proven yet isolated biological knowledge using graph learning methods. This approach reduces the hypotheses from countless possibilities to 8 specific ones to be tested.

While we highlighted one of the potential MoA here, all 153 associations and their potential mechanisms are available in our interactive website.

## Discussion

PregMedNet provides a multifaceted perspective on maternal medication safety and neonatal complications in previously underrepresented populations—pregnant women and neonates. Its contributions include: (a) systematically identifying maternal medication associations with neonatal complications, (b) adjusting for confounding variables across multiple medication-disease pairs, (c) providing *in vivo* experimental support for one of the generated hypotheses, and (d) suggesting potential mechanisms of action (MoA).

The systemic analysis of multiple maternal medications and neonatal complications was conducted by developing mother-baby dyads using retrospective claims data. We advanced our previous algorithms^17^ for linking mother and baby records, estimating gestational age and preterm birth, and incorporating multiple disease features based on International Classification of Diseases (ICD) and Current Procedural Terminology (CPT) codes. While there are previous perinatal drug safety studies using claims databases,^46^ most have focused on short-term outcomes like live birth and stillbirth^17, 19, 20, 47, 48^ or examined a few maternal medication and neonatal complication pairs using traditional statistical testing.^26^ In contrast, we systematically analyzed the impact of all noted maternal medications on 24 different neonatal complications in the cohort, which not only yielded an extensive set of results, but also enabled us to characterize patterns of intricate associations across numerous drug-disease pairs. Additionally, PregMedNet further investigates maternal medication effects at finer resolution by retrieving drug information in a data driven manner at the compound level, and by distinguishing between drug routes of administration. This approach differentiates distinct use cases for the same medication, such as topical betamethasone for skin inflammation and intramuscular betamethasone for preventing complications in preterm labor.

Another key contribution of the study is the adoption of a machine-learning-based confounding adjustment framework, which systematically selects and adjusts confounding variables for each of the 27,648 maternal medication-neonatal complication pairs. This approach aimed to produce more robust associations while eliminating the need for manual selection, making it well-suited for large-scale systematic analyses. The study successfully identified both established and novel associations.

Notably, it reconfirmed previously known associations, such as the increased risk of neonatal respiratory complications in babies perinatally exposed to SSRIs^27^ and the increased risk of NAS in babies exposed to opioids,^49^ demonstrating the capability of the confounding adjustment framework. This framework is expected to scale to a wider range of systematic research studies, including those leveraging claims data or EHR.

Furthermore, the study examined the additive effects of concurrent use of maternal medications on neonatal complications through retrospective DDIs analysis. Notably, the analysis indicated that certain known medication pairs (acetaminophen/oxycodone, NPH/short-acting insulins) may confer a potentially synergistic reduction in risk for neonatal complications. While there have been previous DDI research studies utilizing EHR, these studies have typically concentrated on already known side effects,^50, 51^ being based on data from a single institution,^50, 52^ or mainly focused on a limited number of medication pairs. ^51, 53, 54^ In contrast, we agnostically tested the effects of drug combinations without relying on prior knowledge, using a large-scale claims database from multiple institutions. This approach allows for the discovery of novel interactions and helps ensure more unbiased findings, which is particularly important as polypharmacy among pregnant women has increased over the last three decades.^1^ To the best of our knowledge, this represents the first attempt to screen the effects of maternal drug combinations on neonates based on real-world, retrospective, large-scale claims data from multiple institutions.

Notably, one of the novel findings of PregMedNet—the effect of maternal ondansetron on neonatal BPD— was validated through laboratory experimentation. Our preliminary *in vivo* experiments in mice demonstrated the effects of maternal ondansetron administration on neonatal lung development. Increased mean linear intercept (MLI) could reflect defects in postnatal alveolar septation. Importantly, maternal ondansetron did not alter neonatal survival or weight gain, which can have a secondary effect on lung development.

Ondansetron is a potent and selective antagonist of the 5-HT3A serotonin receptor.^55^ In the developing human lung, the 5-HT3A receptor is expressed on epithelial, endothelial, and smooth muscle cells throughout the canalicular, saccular, and alveolar stages of lung development.^56^ However, the role of serotonin signaling through the 5-HT3A receptor in guiding normal lung morphogenesis is not yet understood. The increased MLI observed in ondansetron-treated mice, as predicted by the PregMedNet findings, suggests that the 5-HT3A receptor may play a role in normal lung morphogenesis.

As such, our *in vivo* experimental validation of a novel medication effect discovered by PregMedNet supports the validity of its predictions. While previous research studies have investigated maternal medication safety using claims databases,^17, 19, 20, 26, 46–48^ to the best of our knowledge, this is the first study to experimentally validate such hypotheses. Notably, this experimentation also generated follow-up hypotheses regarding the previously unknown role of the 5-HT3A receptor in lung development. This further highlights PregMedNet’s potential to guide subsequent hypothesis-driven research. Further investigations into other PregMedNet-generated hypotheses will elucidate mechanisms and identify potential alternatives or treatments.

Understanding a drug’s MoA is critical for developing new treatments and preventing the side effects.^57, 58^ However, traditional approaches to uncovering MoAs are complex and resource-intensive tasks, spanning years, necessitates in-depth hypotheses for testing.^58^ PregMedNet generates hypotheses of MoAs for newly discovered associations that have never been tested before by integrating real-world associations and a knowledge graph. It narrows down the number of molecular targets that need to be tested, potentially leading to significant reduction in time required for further research and validation. A case study with ondansetron and BPD showcased the plausibility of the hypothesis. Although there are previous research studies using graph learning methods to identify drug treatment mechanisms^59^ and polypharmacy drug side effects,^59^ these efforts have relied solely on biological interactions from publicly available databases. PregMedNet distinguishes itself by connecting associations based on a large, real-world patient dataset to a knowledge graph to identify potential MoA. PregMedNet’s ability to suggest potential MoAs is expected to facilitate targeted scientific investigation into underlying biological mechanisms.

Our study was subject to several limitations inherent to the database, the Merative™ Marketscan® Commercial Database. These include the restricted inclusion of patients with private insurance, excluding those without coverage, and the focus on prescribed outpatient medications, which omits inpatient and over-the-counter medications. Critical demographic details such as race, smoking habits, alcohol use, weight, and height are also unavailable. Additionally, there are also limitations inherent to claims data, including challenges in excluding coding errors introduced during billing processes and difficulties in confirming patient compliance with prescribed medication regimens. The limited specificity of ICD and CPT codes presents further challenges, as rare diseases often lack dedicated codes and are grouped under broader categories. For example, in ICD-9, BPD is categorized under code 770.7, described as “Chronic Respiratory Disease arising in the perinatal period,” while in ICD-10, it has a more specific code, P27.1, described as “Bronchopulmonary Dysplasia originating in the perinatal period”. To minimize coding challenges and improve accuracy, three clinicians reviewed the final list of codes after extracting the ICD and CPT codes.

An additional limitation to consider is the complexity and variability of the confounding structure for each association, which may not be fully addressed in the systematic analysis provided by PregMedNet. This may lead to the inclusion of non-causal associations, such as confounding by the indication for the medication and its severity, both of which can vary between associations.

As such, the study has inherent limitations, despite our efforts to maximize the utility of the available data to provide the best possible estimates. To fully establish causal relationships, follow-up analyses aimed at causal inference that incorporate more accurate data along with detailed confounding factors for each association will be necessary. Nonetheless, PregMedNet significantly narrowed the scope to 261 associations involving 68 maternal medications from an initial 27,648 associations, enabling prioritization for further causal investigation. Importantly, our experimental validation of a novel association underscores the reliability of our findings. Taken together, these findings demonstrate PregMedNet’s value as a platform for robust hypothesis generation despite these limitations.

It is also important to consider that medications with statistically non-significant associations in our dataset are not necessarily safe; limited usage of medications already known to pose risks among expectant mothers may reduce the statistical power to assess these associations. We analyzed the frequency of medication use across Pregnancy Categories based on Australian categories for prescribing medicine in pregnancy^60^ (see Supplementary Table 10), which shows that prescription frequency decreases as the risk category increases from A to X. Category X medications (high risk of causing permanent damage to the fetus) were prescribed to only 36 patients, whereas category A medications (drugs without any proven damage to the fetus) were prescribed to 213,019 patients. This distribution supports the notion that higher-risk medications are prescribed less frequently, resulting in less statistical power and explaining why they may not appear in the results. This trend is also reflected in the DDI results, where fewer statistically significant DDIs are observed due to small cohort sizes per drug combination. Thus, non-significant DDIs do not necessarily indicate a lack of effect. However, significant findings in PregMedNet provide risk estimates for commonly used maternal medications, which can further guide decisions on perinatal medication safety.

Finally, while the knowledge graph used for MoA discovery aggregates comprehensive and verified biological associations, it lacks information on the relative strength and directionality of these associations and their specific cellular locations. Including this additional information in future directions may further improve the MoA hypotheses generated through graph learning.

PregMedNet provides a multifaceted approach to studying the impact of perinatal drug use on neonatal outcomes, an area previously underexplored. Using large, multi-institutional claims data and a novel systematic approach, we identified robust associations, one of which was validated through *in vivo* experimentation. PregMedNet also generated hypotheses on potential MoAs for the associations. The extensive results of PregMedNet are available through an interactive website (https://pregmednet.stanford.edu/). We expect PregMedNet will contribute to advancing maternal and neonatal healthcare by providing insights that guide safer medication use during pregnancy and help reduce the risk of neonatal complications.

## Methods

### PregMedNet

PregMedNet is a platform designed to assess the maternal medication impacts during pregnancy and their impact on neonatal complications. PregMedNet provides a comprehensive analysis of the multifaceted associations between 1,152 maternal medications used during pregnancy and 24 neonatal complications, systematically examined retrospectively using large-scale claims data and a novel machine learning method. Notably, it experimentally validated one of the results, demonstrating the reliability of the PregMedNet. Additionally, possible underlying mechanisms of discovered correlations between diseases and medications are identified using Knowledge Graph and graph learning methods. The entire results are open source and accessible through an interactive website (https://pregmednet.stanford.edu/).

#### 1. Data Preparation

Mother and baby cohorts, along with their medical condition and medication histories, were extracted from the large-scale, de-identified claims data of the Merative™ Marketscan® Commercial Database.^21^ This database houses de-identified medical, medication, and dental claim records for over 250 million patients in the US. We accessed the data through the Stanford Center for Population Health Sciences. A conceptual flow diagram of the data preparation process is shown in Supplemental Figure 1 with details in Supplemental Figure 2. The study was reviewed and approved by the international review board (IRB) at Stanford University (no. 39225).

##### 1.a. Determining the Mother and Baby Cohort

The pregnancy cohort was created building upon methodology devised in our previous research involving mother-baby dyads.^17^ We identified a cohort of women aged 12–55 years with singleton pregnancies using the International Classification of Diseases, Ninth Revision (ICD-9), ICD-10, and diagnosis-related group (DRG) codes from 2007-2021 (Supplementary Table 3). We included only the first pregnancies to avoid dependencies among women with multiple pregnancies. Molar and ectopic pregnancies, as well as spontaneous and induced abortions, were excluded (Supplementary Table 4). Only mothers continuously enrolled in the insurance plan up to 90 days before the delivery date were included to ensure all maternal records are captured without any omissions.

The newborn babies’ cohort was generated based on the newborn babies’ codes from ICD-9 and ICD-10 from 2007-2021 (Supplementary Table 5). Only babies continuously enrolled in the insurance plan for 90 days after the delivery date are included to ensure newborn records are captured without any omissions.

##### 1.b. Extracting Relevant Records: Medical Conditions & Medications

For each mother, all medical claims records from 90 days before the delivery date to the delivery date were extracted from both the inpatient and outpatient claims tables. From these records we identified 68 maternal conditions of interest (Supplementary Table 9), defined using ICD codes extracted through either syntax-based keyword searches or manual selection, when required, from publicly available code lists.^61, 62^ ^63^ In addition, we obtained maternal age, gestational age at delivery, preterm birth, and the region and geographic location of the healthcare facility. All maternal medication records during the same period were extracted from the prescription drug claims table as National Drug Codes (NDCs). These medications were further processed as described in the section 1.d.

For each newborn, medical claims records from the delivery date up to 90 days post-delivery were extracted. ICD codes from publicly available lists^62^ ^63^ were used to define 24 neonatal complications (Supplementary Table 8), which were reviewed by three clinicians. Neonatal sex of each baby was also extracted. Maternal data and corresponding baby data were linked using a Family ID, indicating which baby was delivered by which mother (Figure 1.a).

##### 1.c. Estimating Gestational Age and Preterm Birth

Gestational Age (GA) at delivery was determined using GA codes, which are ICD codes that represent the number of completed weeks of gestation (Supplementary Table 6). ICD codes indicating preterm birth (PTB) were designated as PTB codes (Supplementary Table 7). We searched maternal records for both GA and PTB codes spanning one month before to one month after the delivery date. This wide temporal window was necessary to account for the occasionally inaccurate timing of claims data.

GA estimation proceeded as follows:

1. If a single GA code was recorded in maternal records, that value was extracted as the GA. If multiple GA values are recorded, the highest recorded GA value was selected.
2. If no GA code was recorded in maternal records, but infants had a GA code in their records within one month of delivery, GA was inferred from the infant record.
3. If no GA code was recorded in both maternal and infant records, but PTB codes were recorded, a GA of 34 weeks was assigned. Otherwise, GA of 39 weeks was designated.

Preterm birth records were estimated similarly:

1. If PTB codes were recorded in maternal records, the pregnancy was associated with PTB.
2. If no PTB codes were recorded in maternal records, but GA code suggested less than 37 weeks of gestation, the pregnancy was considered to be delivered preterm. Otherwise, a full-term delivery was assumed.
3. If mothers had both PTB codes and GA codes, but GA was higher than 37 weeks, they were removed from the cohort.

##### 1.d. Maternal Medications

Generic drug names, drug classes, and route of administration information from MarketScan Redbook^64^ were additionally integrated to the maternal medication records using NDC. Medication records were further processed as follows (Supplementary Figure 3):

1. Categorized medication claims records by their route of administration, identifying 24 distinct routes.
2. For each route of administration, generic medication records were split at the compound level using syntax-based parsing of slashes and semi-colons.
3. Next, medication names were standardized using DrugBank IDs^65^ to account for compounds described differently (e.g., Oxycodone Hydrochloride vs. Oxycodone HCl). Medications without DrugBank IDs, totaling 139, were manually reconciled. Out of these, 31 medications were assigned manual IDs, while the rest were discarded.
4. Each final medication consisted of a harmonized medication name and route of administration such that the same medication taken by different routes of administration was considered separately. Medication administration was binarized so that each medication was designated as “taken” or “not taken”.

##### 1.e. Final Mother-Baby Cohort

The final dataset included 1,152 maternal medications and 68 maternal covariates during the 90 days leading up to delivery, as well as data on 24 neonatal complications during the first 90 days post-delivery. It also included 6 additional attributes: maternal age, gestational age at delivery, preterm birth, baby’s sex, and the region and geographic location of the healthcare facility. Except for the demographic details, all other variables in the dataset were binary variables to indicate the presence or absence of a condition or medication.

The final dataset was split into a discovery cohort and validation cohort. Babies delivered between 2007 and 2015 were included in the discovery cohort, while those delivered between 2016 and 2021 were used for validation.

#### 2. Odds Ratios: Maternal Medications and Neonatal Complications

The associations between maternal medications during pregnancy and neonatal complications were calculated using both unadjusted and adjusted odds ratios. Odds ratios were derived from logistic regression, while adjusted odds ratios were determined through Lasso Logistic Regression Without Penalizing Exposure Variable (LassoNoExp).^66, 67^ Potential confounders were selected using variance inflation factors (VIF) from among 68 maternal comorbidities and 6 additional attributes (maternal age, gestational age at delivery, preterm birth, and the region and geographic location, the sex of each baby) to mitigate the effects of multicollinearity. We next describe this approach in detail.

##### 2.a. Odds Ratios

Odds ratios (ORs) were computed using univariate logistic regression for each medication-disease pair. In equation (1), 𝑌 ∈ {*0*,*1*}^817,402^, represented the neonatal complication vector, and 𝑀 ∈ {*0*,*1*}^817,402^, denoted the medication vector where entry *𝑀_j_* equaled 1 if the mother indexed by 𝑗 was exposed to the medication, and 0 otherwise.Therefore, the odds ratio for each pair was given by equation (2). This process was repeated for all possible pairs, yielding 27,648 ORs from 1,152 medications and 24 diseases. The Benjamini-Hochberg procedure^68^ was subsequently applied to control the false discovery rate (FDR) at 5% to correct for multiple comparisons.

- Logistic Regressions between each medication and neonatal complication pair

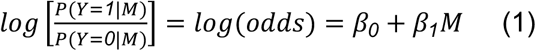

- Odds Ratio

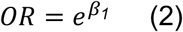

##### 2.b. Adjusted Odds Ratios

While ORs from 2.a. provided an estimate of the effects of maternal medication, they could be influenced by spurious associations due to unaccounted confounders. However, traditional methods of manually selecting confounders were impractical for our study of 27,648 associations, and uniformly adjusting for high-dimensional confounders also posed a challenge due to overadjustment bias^69^ and multicollinearity^70^. To obtain a more accurate representation of the maternal medication’s impact on neonatal complications, we adjusted the odds ratios for 74 selected potential confounders. These adjusted odds ratios (aORs) were computed using Lasso Logistic Regression Without Penalizing Exposure Variable (LassoNoExp) after removing certain covariates to mitigate multicollinearity.

###### 2.b.1. Managing Multicollinearity of Confounders

Multicollinearity refers to correlation of independent variables in multiple regression analysis. This poses challenges for both mathematical modeling and results interpretation. For a given multiple regression 𝑦 = 𝑋𝛽 + 𝜖, the ordinary least squares (OLS) is given by *β̂* = (𝑋^𝑇^𝑋)^−^*^1^*𝑋^𝑇^𝑦. However, if columns of 𝑋 are linearly dependent and therefore correlated, then 𝑋^0^𝑋 becomes non-invertible (singular), making it impossible to complete the OLS formula. Highly correlated columns are also difficult to interpret because it becomes difficult to discern the individual contributions of each factor to predicting the dependent variable, and increases the variance of the regression coefficient causing it to become unstable.^71^

In our study, we initially included a total of k=74 potential confounders, encompassing maternal comorbidities and demographic variables, identified based on previous research related to maternal morbidity.^72^ To reduce multicollinearity, we first selected independent variables (i.e. confounders) based on the Variance Inflation Factor (VIF).^73^ VIF is a measure of multicollinearity that indicates the degree of intercorrelation among the independent variables. VIF of each confounder was calculated by fitting a linear regression model using one confounder as the dependent variable and all others as independent variables (3). Subsequently, the multiple correlation coefficient of 𝑖^th^ confounder 𝑋_1_, 𝑅_1_*^2^*, was used to calculate VIF of 𝑖^th^ confounder 𝑋_1_, as given by (4). This process was repeated for all confounders, with a higher VIF indicating greater multicollinearity.

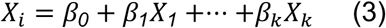

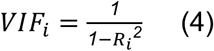

In our model, we excluded six confounders that had either a VIF greater than 5^74^ or zero vectors. This exclusion included specific substance use disorders (e.g., fentanyl, amphetamine, ecstasy, methamphetamine, methylphenidate) and region of the healthcare facility features. Nonetheless, we retained both gestational age and maternal age, despite their VIFs exceeding 5, due to their critical role in determining outcomes. After this process, a total of 68 confounders remained, which were used for adjustment.

###### 2.b.2. Feature Engineering: Demographic Features

Maternal age, gestational age, and the geographic location of the healthcare facility features were further processed. Because maternal age and gestational age had a wider span of values compared to binary inputs which could have led to them being overemphasized in the model, they were scaled using a min-max scaler. The geographic location of the healthcare facility was a categorical variable with 54 categories, which can introduce additional multicollinearity. Thus, it was encoded using a binary encoding method, expressing 54 variables in 6 columns. The final dataset for adjusted odds ratios contained 73 columns representing confounders, along with 1,152 maternal medications and 24 neonatal complications.

###### 2.b.3. Additional Adjustment for Postmaturity

When the outcome variable 𝑦 was postmaturity, preterm birth and gestational age were excluded from the confounders due to their potential to introduce multicollinearity, as both were defined by gestational age. As a result, the total number of confounders for postmaturity was reduced to 66, compared to 68 for the other 23 neonatal complications. This adjustment led to a final dataset containing 71 columns representing confounders, along with 1,152 maternal medications, specific to postmaturity, distinguishing it from the datasets used for the other neonatal complications.

###### 2.b.4. LASSONoExp: Adjusted Odds Ratios Calculation

We adjusted the confounders using LASSONoExp, an algorithm based on LASSO regression.^75, 76^ Lasso regression is effective for adjusting potential confounders and estimating treatment effects due to its ability to select variables among high-dimensional confounders and reduce overfitting.^77^ LASSONoExp is the same as the LASSO, except that the regression coefficient of the exposure variable, in our case, maternal medication exposure, is not penalized in the model. This prevents the coefficient of maternal medication from yielding a zero value. Consequently, the exposure effect of the estimator is not biased, while the effects of the confounding variables are regularized.^66, 67^

Let 𝑀 denote the maternal medication of interest, as defined in Equation (1). As before, 𝑌 represents the neonatal complications of interest, and 𝑋_3_(where *2* ≤ 𝑖 ≤ 𝑃) indicates the confounders that were selected from section 2.b.1 to 2.b.3 (in our case, 𝑃 = *71* for postmaturity, and 𝑃 = *73* for the rest of the neonatal complications) such that 𝑋*_ij_* = *1* if the mother indexed by 𝑗 had comorbidity 𝑖 and 0 otherwise. The model is then given by

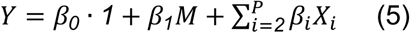

Coefficients 𝛽*_1_* are determined by

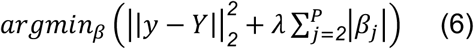

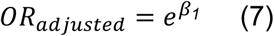

In LASSONoExp, the coefficients β_i_, as denoted in (5), are obtained by solving the objective function (6) where hyperparameter γ represents a penalty weight for the confounders. In (6), the term *y* represents the ground truth (actual observed values), whereas *Y* signifies the predicted values obtained from the model. Here, β_1_, the coefficient of maternal medication, is not penalized, while β_2_ to *β_P_* are penalized based on the penalty weight. Adjusted odds ratios are then given by (7).

We repeated this process for the 1,446 maternal medication and neonatal complication pairs that yielded statistically significant unadjusted odds ratios, as described in section 2.a. The Python package *statsmodels* was used to fit the model. The penalty weight, *λ*, was selected for each maternal medication-neonatal complication pair based on 3-fold stratified cross-validation. This selection involved 10 coefficients ranging between 0.01 and 10, using the Python package *optum*. Finally, the Benjamini-Hochberg procedure was applied to control the FDR at 5%, correcting for multiple hypothesis testing.

##### 2.c. Network Graph

Unadjusted odds ratios and adjusted odds ratios, given by equations (2) and (7) respectively, were visualized as a network graph. This visualization was chosen to illustrate the pattern of interactions and to allow exploration of multiple correlations in a single plot, given that there are more than 1,000 statistically significant correlations. (See Figure 2, Supplementary Figure 4, and the interactive graph at http://pregmednet.stanford.edu). In the figures, each node represents either neonatal complications or maternal medications, while the edges between them indicate statistically significant odds ratios. The size of the nodes is proportional to the number of edges connected to them, and the width of the edges is inversely proportional to the p-values of the odds ratios. The color of the nodes represents either the medication group or disease (with gray-colored nodes at the center), and the color of the edges indicates whether the odds ratios are positive (gray) or negative (blue). The locations of the nodes were estimated based on t-distributed Stochastic Neighbor Embeddings (t-SNE)^78^ within each disease or medication group, demonstrating relative similarities in the patterns of medication use or disease outbreaks in mother-baby dyads. The Python package *scikit-learn* was used to calculate t-SNE, *networkx* to generate the network graph, and *bokeh* for creating an interactive graph.

##### 2.d. Verification of Results in the Validation Cohort

To ensure the reproducibility of the odds ratios and adjusted odds ratios, we repeated the same process on the validation cohort and compared the results of unadjusted and adjusted odds ratios between the two cohorts by matching maternal medications and neonatal complications. After matching, scatter plots were created, and Pearson correlation coefficients were calculated between discovery and validation cohort odds ratios.

#### 3. Drug-Drug Interactions (DDIs)

##### 3.a. Derivation of DDIs

Drug-Drug Interactions (DDIs) occur when two or more medications react with each other affecting the outcome in a way that would otherwise not occur. DDIs are considered in pairs of maternal medications relative to each of the 24 neonatal complications. This is done to determine whether interactions between two medications could additionally affect neonatal complications. Gestational Age, one of the strongest confounders, is included and adjusted for in the analysis. DDI effects were captured using a logistic regression-based model,^79^ given by:

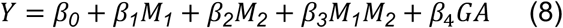

where GA denotes gestational age, 𝑀_3_ (𝑖 = *1*,*2*) denotes exposure to maternal medication 𝑖, and 𝑌 represents the neonatal complication of interest. 𝑀*_1_*𝑀*_2_* represents the interaction between medication 1 and medication 2, and its coefficient, 𝛽*_3_*, indicates the direction and magnitude of this interaction. If 𝛽*_3_* is less than 0, it suggests that the interaction tends to reduce the risk of neonatal complications. Conversely, a 𝛽*_3_* greater than 0 indicates that the interaction increases the risk of neonatal complications. The higher absolute value of 𝛽*_3_* suggests stronger interactions.

We repeated this process for all maternal medication pairs among 1,152 medications for 24 neonatal complications. We only considered interactions where there were more than 10 mother-baby dyads where the mothers took both medications and the babies had neonatal complications. A total of 28,417 maternal medication pairs and neonatal complications out of the 15,911,424 all possible pairs (calculated as 1152*C_2_* × 24) were analyzed. The Benjamini-Hochberg procedure was then applied to control the FDR at 5%, correcting for multiple hypothesis testing. Ultimately, a total of 121 maternal medication pairs and neonatal complications showed statistically significant 𝛽*_3_*.

##### 3.b. Calculate the Odds of Drug Pairs

For the 121 maternal medication pairs and neonatal complications that exhibited statistically significant 𝛽*_3_* values computed from Section 3.a., odds ratios for concomitant medication use were calculated as:

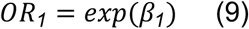

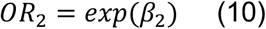

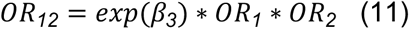

As with 𝛽_3_, the Benjamini-Hochberg correction was subsequently applied to 𝛽_1_ and 𝛽_2_, respectively, to control the FDR at 5%. Equation (11) was applied when all 𝛽_3_, 𝑂𝑅_1_, 𝑂𝑅_2_ are statistically significant. Finally, we extracted the medication pairs that showed statistically significant 𝑂𝑅*_12_* among the 121 medication pairs, and then compared these results with the odds ratios of their corresponding single medication use cases.

##### 3.c. Final DDIs Overlap: Discovery vs. Validation Cohorts

The same process was repeated within the validation cohort to identify significant values of 𝛽*_3_* and 𝑂𝑅*_12_*. DDIs for which both 𝛽*_3_* and 𝑂𝑅*_12_* were found to be significant in both the discovery and validation cohorts were extracted for further analysis.

#### 4. *In vivo* ondansetron experiments

The association between maternal ondansetron and neonatal bronchopulmonary dysplasia, one of the associations identified in section 2, is tested experimentally.

##### 4.a. Mice

Female and male C57BL/6 mice were purchased from the Jackson Laboratory and maintained in-house. All animal experiments were approved by Stanford University’s Institutional Animal Care and Use Committee under protocol 33897.

##### 4.b. Ondansetron administration

Ondansetron (Millipore Sigma 1478571) was dissolved in PBS without calcium and magnesium (Gibco 10010-049). C57BL/6 female and male mice were timed mated and pregnancy was confirmed by a 2 gram increase in weight by E10 ^80^. Starting on E15, pregnant mice were dosed daily with either 5 mg/kg body weight of ondansetron or PBS by intraperitoneal injection until delivery of pups.

##### 4.c. Lung collection for histology

On P11, pups were euthanized and the lungs were inflated with 4% PFA (Electron Microscopy Sciences 15713). Following inflation, the lungs were collected and fixed in 4% PFA for 1 hour at room temperature. The lungs were then transferred to 15% sucrose (Millipore Sigma S0389) and incubated overnight at 4°C. The following day, the lungs were transferred to 30% sucrose and incubated for 3 days at 4°C, followed by embedding in OCT mounting medium (Leica 14020108926) for cryo-sectioning. Lungs were sectioned at a thickness of 10μm.

##### 4.d. Hematoxylin and eosin staining

For hematoxylin and eosin staining, slides were brought to room temperature and fixed in 10% buffered formalin (StatLab FXBFOLT) for 20 minutes. Slides were immersed in Gill II hematoxylin (Newcomer Supply 1180D) for 15 seconds, rinsed with water, and immersed in Scott’s bluing reagent (Ricca Chemical Company 66971) for 30 seconds. Slides were then rinsed with water, immersed in eosin Y (Millipore Sigma HT110116) for 2 minutes, and again rinsed with water. Slides were dehydrated with dips in ethanol (Fisher A962P-4) and cleared with dips in xylene (Fisher X5-4) before being mounted with non-aqueous mounting medium (Vector Laboratories H570060).

##### 4.e. Mean linear intercept quantification

For quantification, brightfield images were captured with the Leica Thunder microscope using the 20X objective. Images were quantified on FIJI using the Mean Linear Intercept plug-in ^81^. The image analysis parameter was set to 500 pixels between lines, and both horizontal and vertical chords were measured. All horizontal and vertical chords were averaged to get an image average value. For each pup, 3 representative lung sections were stained and imaged, with 5 images taken from each section for a total of 15 images quantified.

#### 5. Knowledge Graph: Hypothesis Generation for New Medication Effects

The hypotheses of underlying mechanisms for newly discovered medication effects were explored using the Precision Medicine Knowledge Graph (PrimeKG).^82^ This publicly available knowledge graph integrates 20 different biological databases, providing a holistic view of the interactions among 10 different types of biological entities. In PrimeKG, biological entities are represented as nodes, and their interactions as edges. Using this framework, we tested two different types of subgraph configurations.

##### 5.a. Subgraph Configuration 1

First, we extracted a subgraph consisting of four biological node types: proteins/genes, biological processes, molecular functions, and cellular components. The subgraph included seven types of edges representing various interactions: protein-protein, bioprocess-protein, molfunc-protein, cellcomp-protein, bioprocess-bioprocess, molfunc-molfunc, and cellcomp-cellcomp.

Next, we incorporated the disease node of interest and its connections to proteins through disease-protein edges. When the disease node was unavailable, we replaced it with an effect/phenotype node.

To ensure the subgraph remained fully connected and did not contain isolated components, we iteratively expanded it by adding additional biological node types (phenotype, disease_phenotype_positive, disease_phenotype_negative) as needed. This expansion was performed per disease type, ensuring that all relevant nodes and edges formed a single, connected network.

We then incorporated the medication node of interest, linking it to proteins through drug-protein edges. Once the subgraph was fully constructed, we extracted all possible shortest paths between the two key nodes of interest—a maternal medication and a neonatal complication—using the Dijkstra algorithm.^83^ The resulting list of shortest paths was considered indicative of potential underlying mechanisms, providing insights into how maternal medications might contribute to neonatal complications.

##### 5.b. Subgraph Configuration 2

For comparison, we conducted the same analysis using a simplified subgraph consisting of only three node types—drugs, proteins/genes, and diseases—and three edge types (drug-protein, protein-protein, and disease-protein). As before, we extracted all possible shortest paths between the maternal medication and the neonatal complication to infer potential mechanisms.

##### 5.c. Iterative Analysis & Visualization

This process was repeated for all newly discovered associations between maternal medications and neonatal complications that exhibited statistically significant adjusted odds ratios (as described in section 2.b). The resulting hypotheses were visualized as graphs using the Python packages *networkx* and *bokeh*.

## Data Availability

The Merative Marketscan Commercial Database is available to purchase by federal, nonprofit, academic, pharmaceutical, and other researchers. Use of the data is contingent on completing a data use agreement and purchasing the data needed to support the study. More information about licensing the Merative Marketscan Commercial Database is available at: https://www.merative.com/documents/brief/marketscan-explainer-general.

## Data Availability

The Merative™ Marketscan® Commercial Database is available to purchase by federal, nonprofit, academic, pharmaceutical, and other researchers. Use of the data is contingent on completing a data use agreement and purchasing the data needed to support the study. More information about licensing the Merative™ Marketscan® Commercial Database is available at: https://www.merative.com/documents/brief/marketscan-explainer-general.

## Code Availability

The PregMedNet framework and custom computer code used in this study can be accessed on GitHub (https://github.com/yerakim824/PregMedNet).

## Author Contribution

Y.K., I.M., G.M.S., D.K.S., and N.A. conceived the study. Y.K., I.M., G.M.S., B.T.B., D.K.S., L.S.P., and N.A. conceptualized and designed the study. Y.K., I.M., and G.M.S. collected and investigated claims data. Y.K. processed the claims data. Y.K. and L.H. developed and reviewed the medication cleanup method. Y.K., J.D.R., and W.M.K. extracted and reviewed the ICD codes. Y.K., I.M., and N.A. developed the computational methodology. Y.K. conducted computational experiments. Y.K. developed the software. L.D.B. and L.S.P. designed the in vivo experiments. C.M.K. performed ondansetron administration. C.M.K., L.D.B., and K.J.C. harvested lung tissue. C.M.K. sectioned, stained, and imaged lungs for mean linear intercept quantification. C.M.K. performed statistical analyses for the in vivo experiments. Y.K. wrote the original draft. C.M.K. wrote the in vivo experimentation section of the manuscript. I.M., P.C., J.D.R., L.S.P. and N.A. reviewed the original manuscript. All authors reviewed and edited the final manuscript.

## Ethics Declaration

### Competing Interests

N.A. is a cofounder of Takeoff AI, a member of the scientific advisory boards of January AI, Parallel Bio, Celine Therapeutics, and WellSim Biomedical Technologies, and a paid consultant for MaraBio Systems. B.G. is an advisory board member at SurgeCare. D.K.S. is a member of the Clinical Advisory Board of Maternica Therapeutics. The remaining authors declare no competing interests.

## Acknowledgement

Data for this project were accessed using the Stanford Center for Population Health Sciences Data Core. The PHS Data Core is supported by a National Institutes of Health National Center for Advancing Translational Science Clinical and Translational Science Award (UL1TR003142) and from Internal Stanford funding. The content is solely the responsibility of the authors and does not necessarily represent the official views of the NIH. Certain data were supplied by Merative as part of one or more MarketScan Research Databases. Any analysis, interpretation, or conclusion based on these data is solely that of the authors and not Merative.

---

**Supplementary Figure 1.**
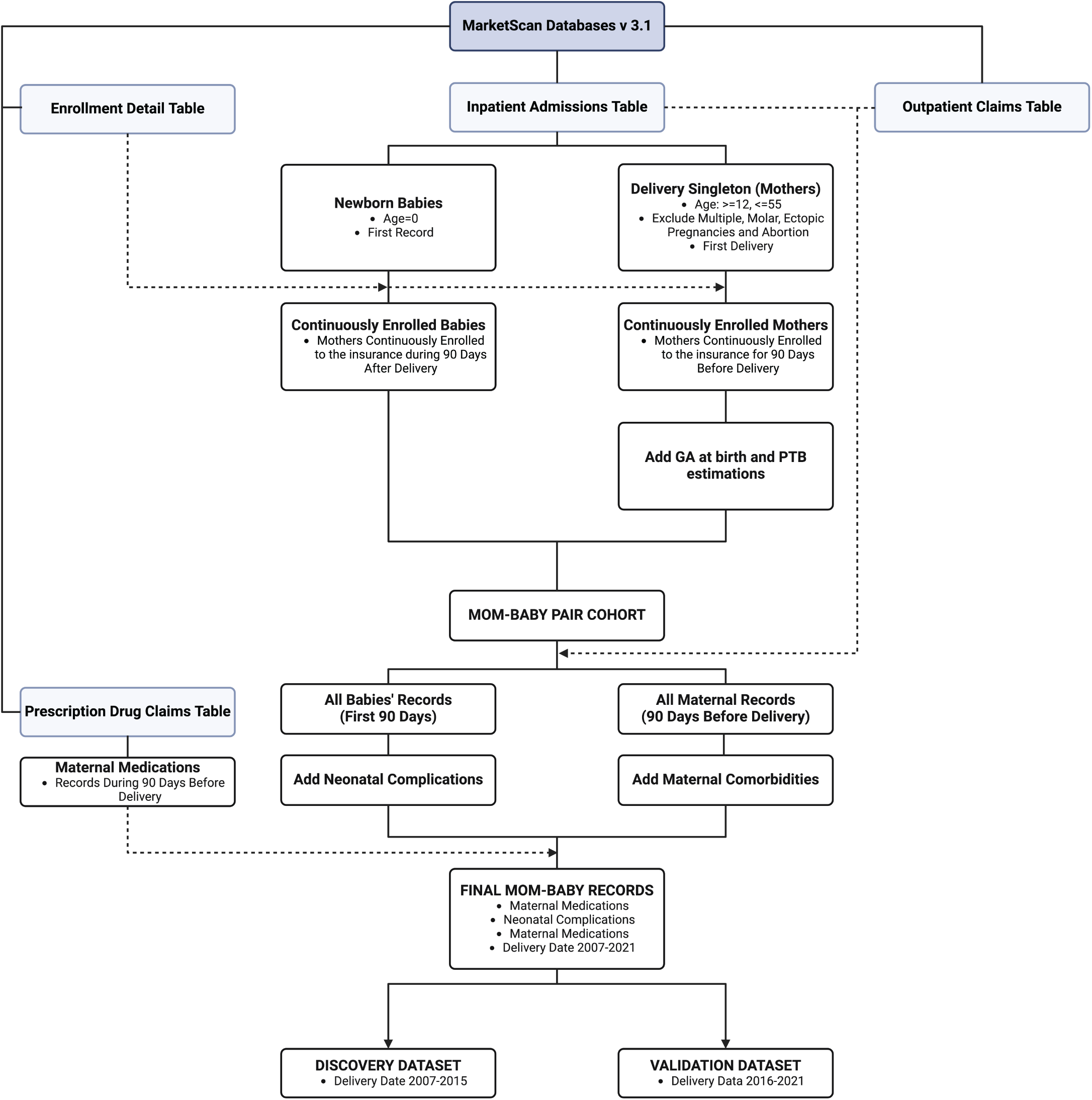
Flow Chart of the Data Construction Overview. This figure illustrates the high-level process of data cleaning and preparation for analysis derived from the Merative™ Marketscan® Commercial Database.

**Supplementary Figure 2.**
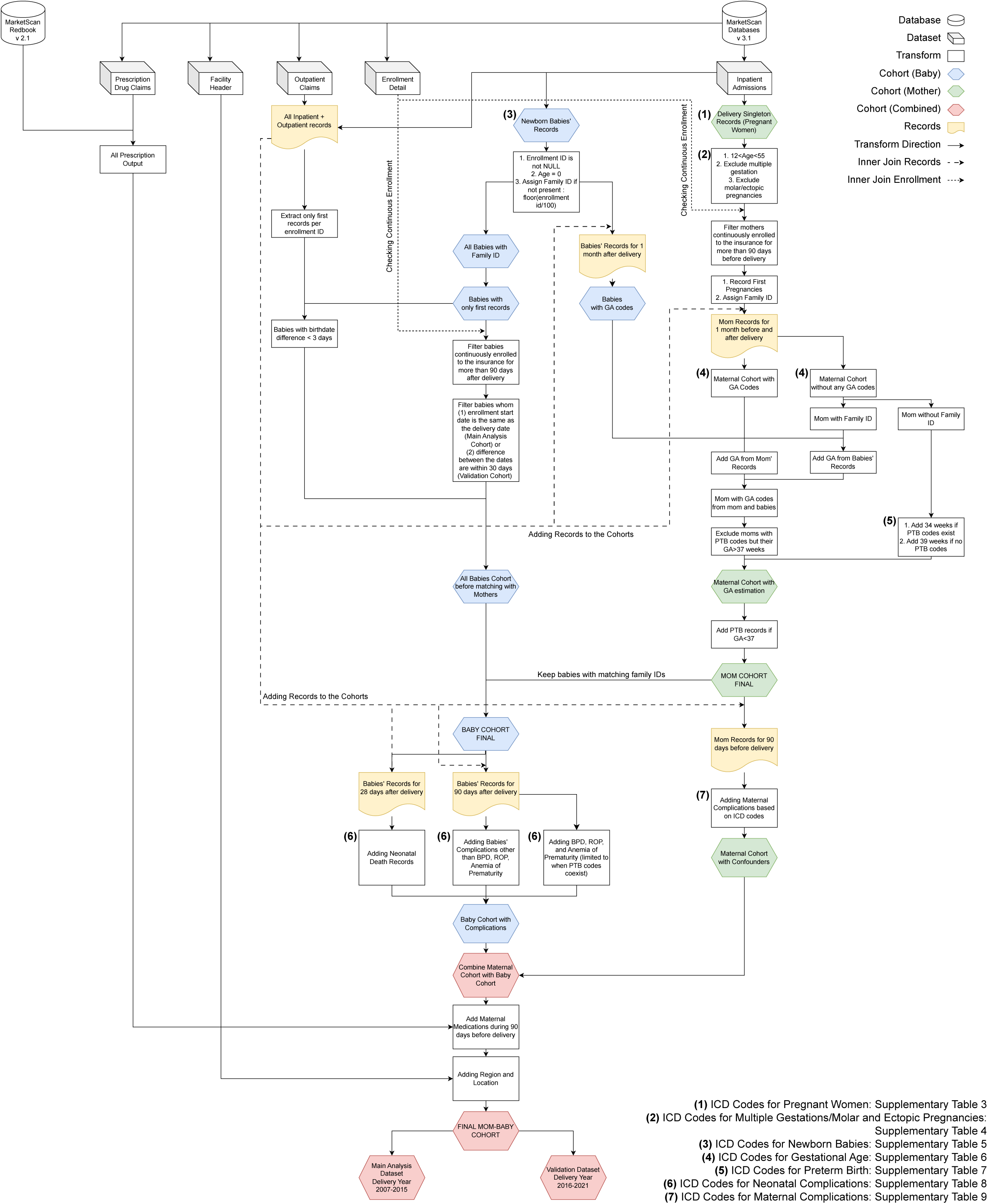
Comprehensive Flowchart Detailing Data Construction. This figure details the extensive process of data cleaning and preparation undertaken using the Merative™ Marketscan® Commercial Database. It expands upon the methodologies outlined in Supplementary Figure 1, providing clarity on the data preparation steps for the study. The flowchart depicts the procedural actions executed to assemble and refine the dataset, leading to the creation of a table that encapsulates maternal medication use, maternal comorbidities, 6 other attributes (maternal age, gestational age at delivery, preterm birth, baby’s gender, and the region and geographic location of the healthcare facility), and neonatal complications. This offers a thorough perspective at the individual patient level and serves to enhance the foundational information displayed in Figure 1(a).

**Supplementary Figure 3.**
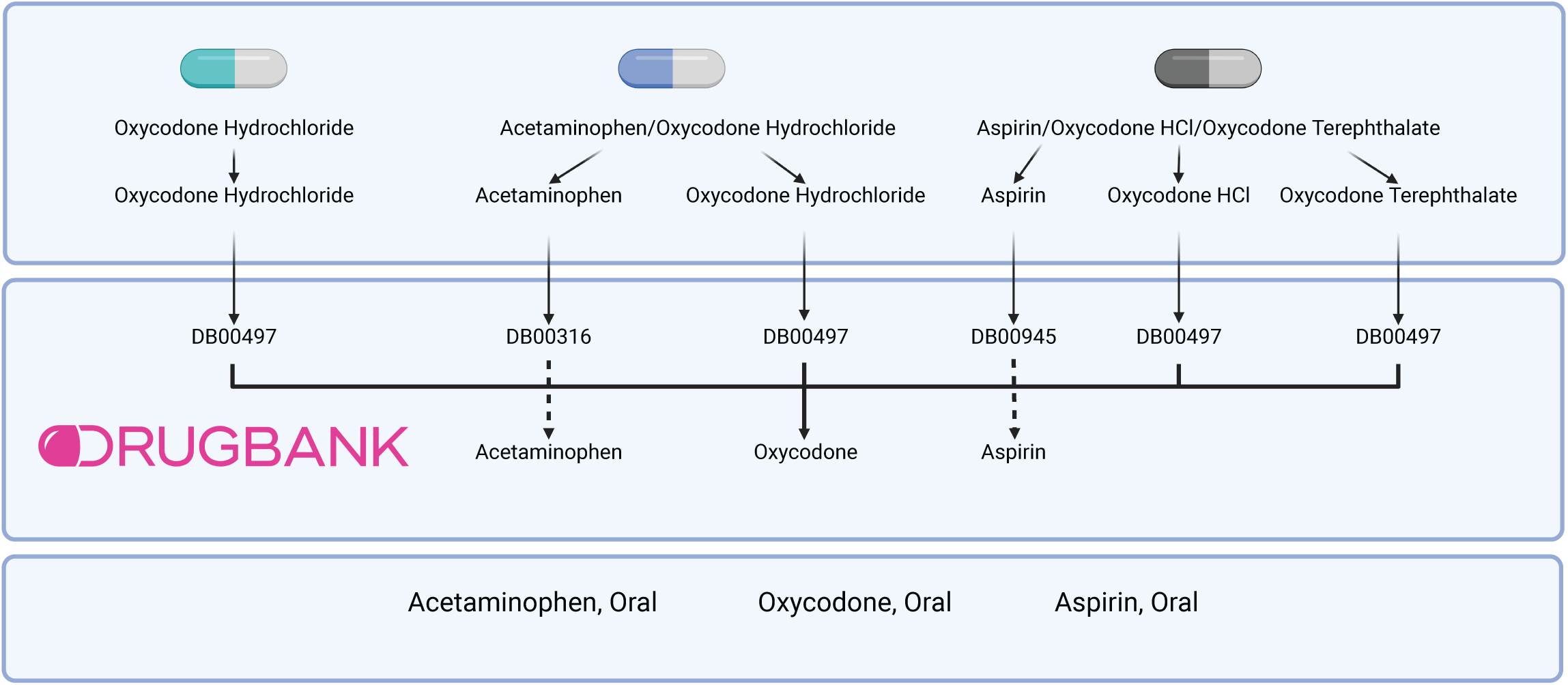
Medication Name Harmonization. This figure outlines the process employed to clean up and harmonized maternal medication data. 1) Maternal medication with generic names was divided at the compound level. 2) Identical medications with varying names were consolidated and harmonized by employing DrugBank IDs along with their common names. 3) Lastly, information regarding the route of administration was integrated into these standardized names.

**Supplementary Figure 4.**
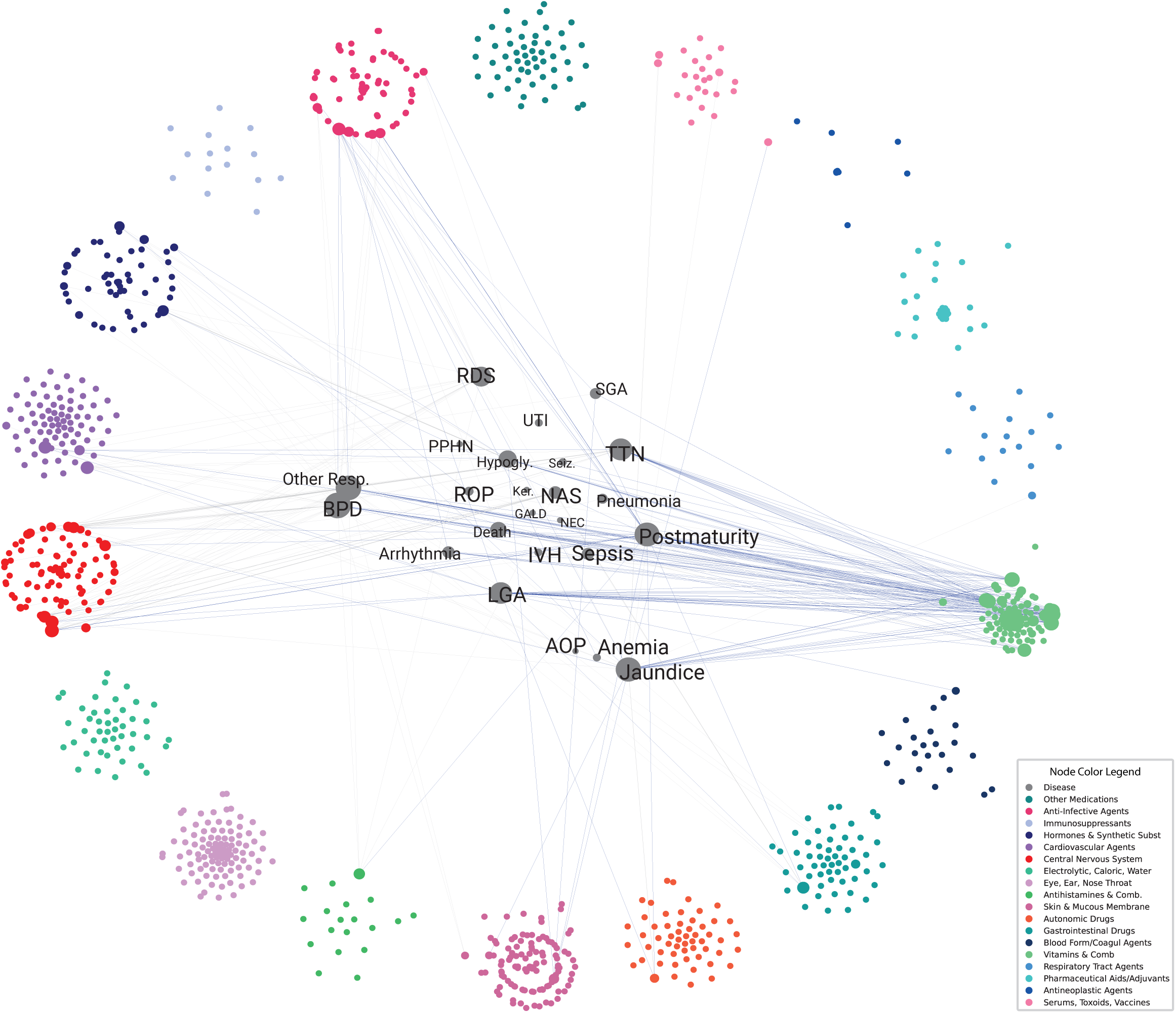
Network Graph of the Adjusted Odds Ratios Between Maternal Medications and Neonatal Complications. Network graph depiction of adjusted odds ratios (aORs) between maternal medications and neonatal complications. aORs are calculated using LassoNoExp, a method that selects confounders by penalizing the excess of potential confounders for each medication-disease pair. Each node in the network graph represents either a maternal medication or a neonatal complication, consistent with the representation in Figure 2. Edges between nodes indicate statistically significant aORs, differentiated from unadjusted odds ratios presented Figure 2. PregMedNet identifies 261 statistically significant adjusted odds ratios (aORs) after adjustment, involving the associations of 68 maternal medications and 19 neonatal complications. Out of the total 261 edges, the 117 edges representing aORs > 1, indicating an increased risk association, are shown as gray edges, while the 144 edges representing aORs < 1, indicating a reduced risk association, are colored blue. The abbreviations for disease nodes are as defined in Figure 2.

**Supplementary Figure 5.**
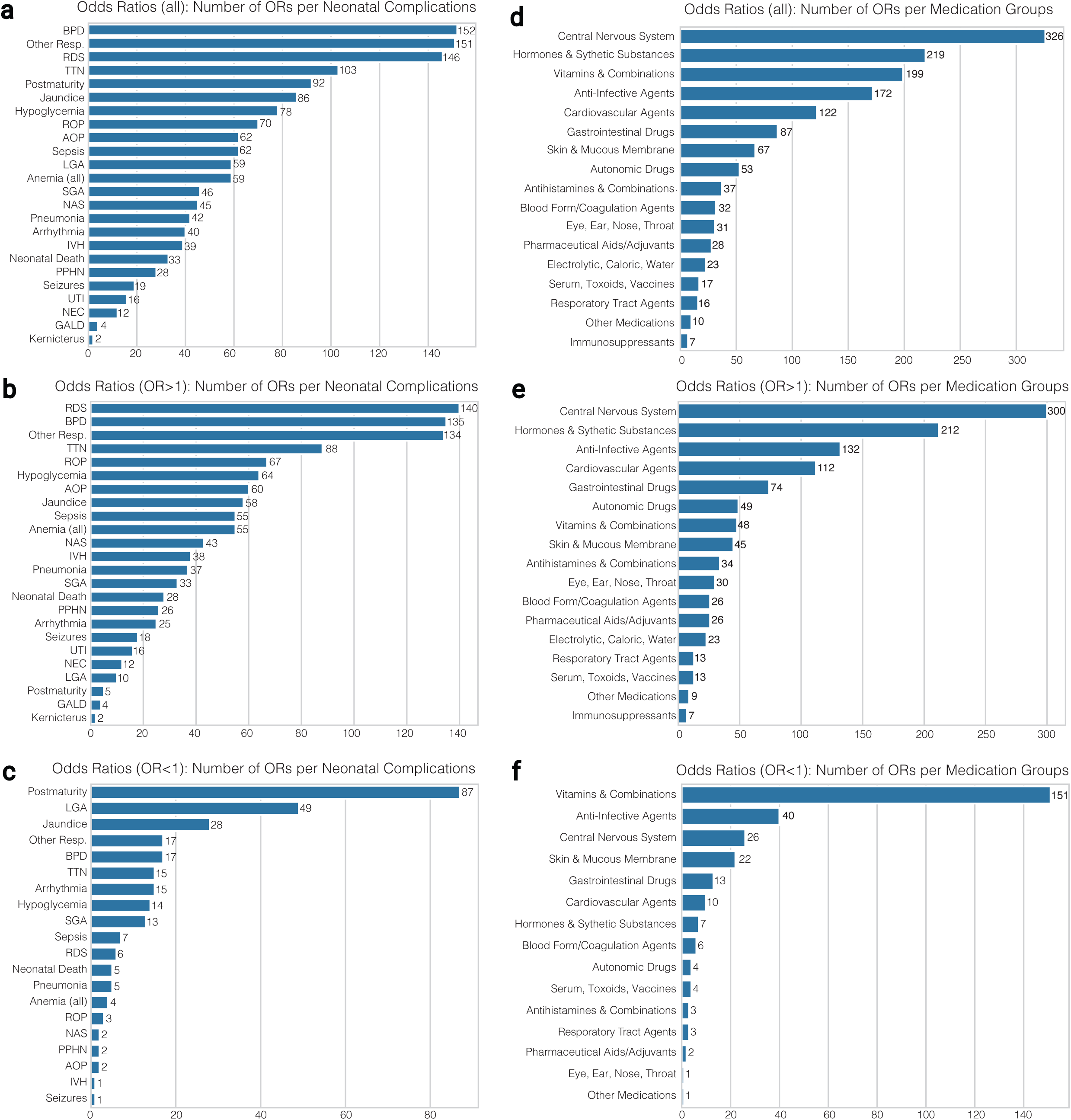
Distribution of Unadjusted Odd Ratios for each Neonatal Complications and Maternal Medicaton Group. Bar graphs that show the count of statistically significant unadjusted odds ratios (OR). (a) The number of all statistically significant odds ratios associated with neonatal complications. BPD showed the most significant associations, followed by other respiratory disorders, RDS, TTN, and Postmaturity. (b) The number of statistically significant increased risk odds ratios associated with neonatal complications. RDS showed the most increased risk associations, followed by BPD, other respiratory disorders, TTN, and ROP. (c) The number of statistically significant reduced risk ORs associated with neonatal complications. Postmaturity has the most reduced risk edges followed by LGA and Jaundice. (d) The number of all significant ORs categorized by maternal medication groups. Medications in the Central Nervous System showed the most significant associations, followed by Hormones & Synthetic substances, Vitamins & Combinations, and Anti-Infective Agents. (e) The number of significant increased risk ORs categorized by maternal medication groups. Medications in the Central Nervous Systems show the most increased risk associations, followed by Hormones & Synthetic Substances, Anti-Infective Agents, and Cardiovascular Agents. (f) The number of significant reduced risk ORs categorized by maternal medication groups. The Vitamins & Combinations group shows the most reduced risk associations, followed by Anti-Infective Agents.

**Supplementary Figure 6.**
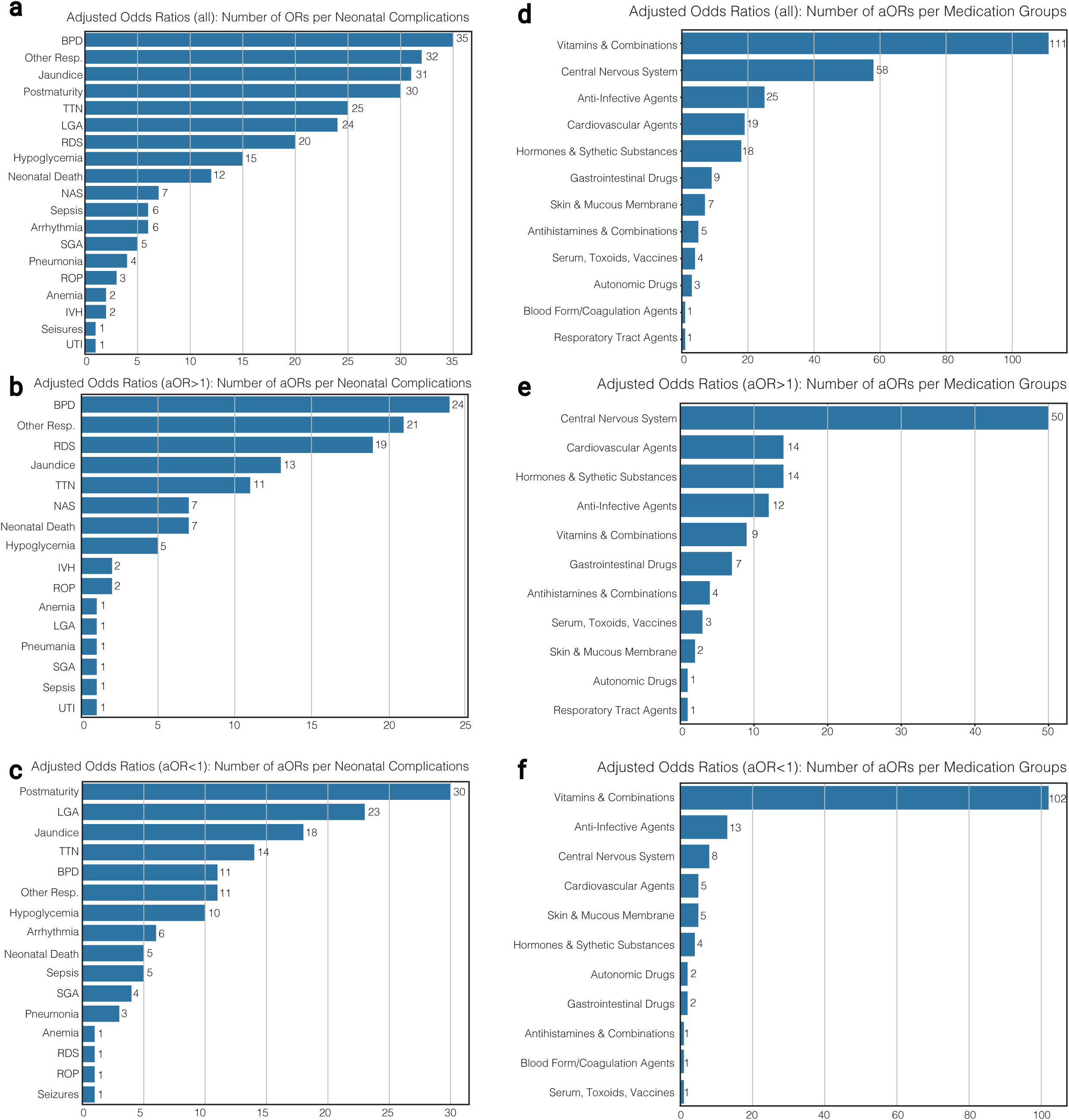
Distribution of Adjusted Odd Ratios for each Neonatal Complications and Maternal Medicaton Group Bar graphs that show the count of statistically significant adjusted odds ratios (aOR). (a) The number of all statistically significant aORs associated with neonatal complications. BPD showed the most significant associations, followed by other respiratory disorders, Jaundice, Postmaturity, and TTN. (b) The number of statistically significant increased risk aORs associated with neonatal complications. BPD showed the most increased risk associations, followed by other respiratory disorders, RDS, Jaundice, and TTN. (c) The number of statistically significant reduced risk aORs associated with neonatal complications. Postmaturity has the most reduced risk edges followed by LGA and Jaundice. (d) The number of all significant aORs categorized by maternal medication groups. Medications in the Vitamins & Combinations showed the most significant associations, followed by Central Nervous System, and Anti-Infective Agents. (e) The number of significant increased risk aORs categorized by maternal medication groups. Medications in the Central Nervous Systems show the most increased risk associations, followed by Cardiovascular Agents, Hormones & Synthetic Substances, and Anti-Infective Agents. (f) The number of significant reduced risk aORs categorized by maternal medication groups. The Vitamins & Combinations group shows the most reduced risk associations, followed by Anti-Infective Agents.

**Supplementary Figure 7.**
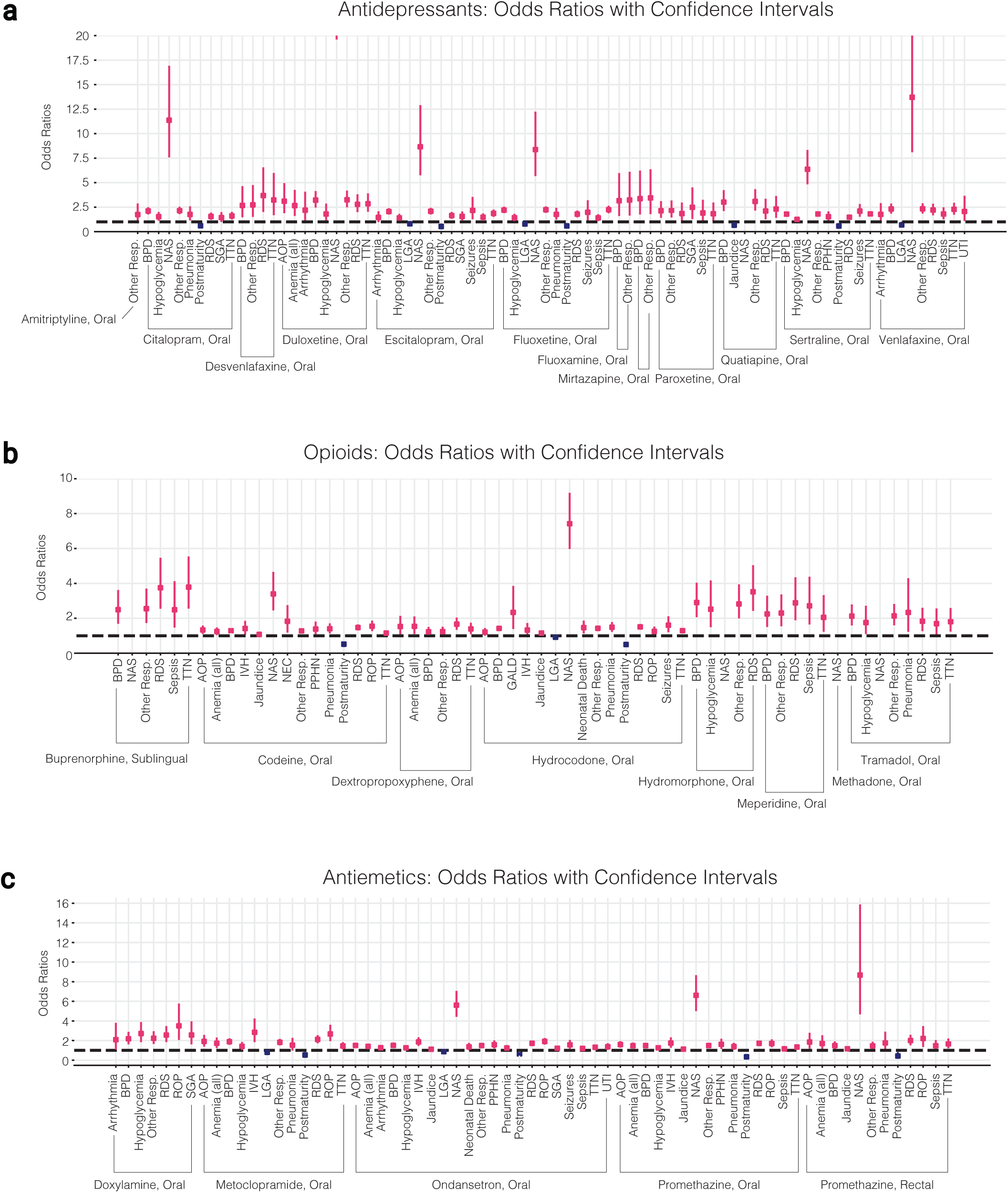
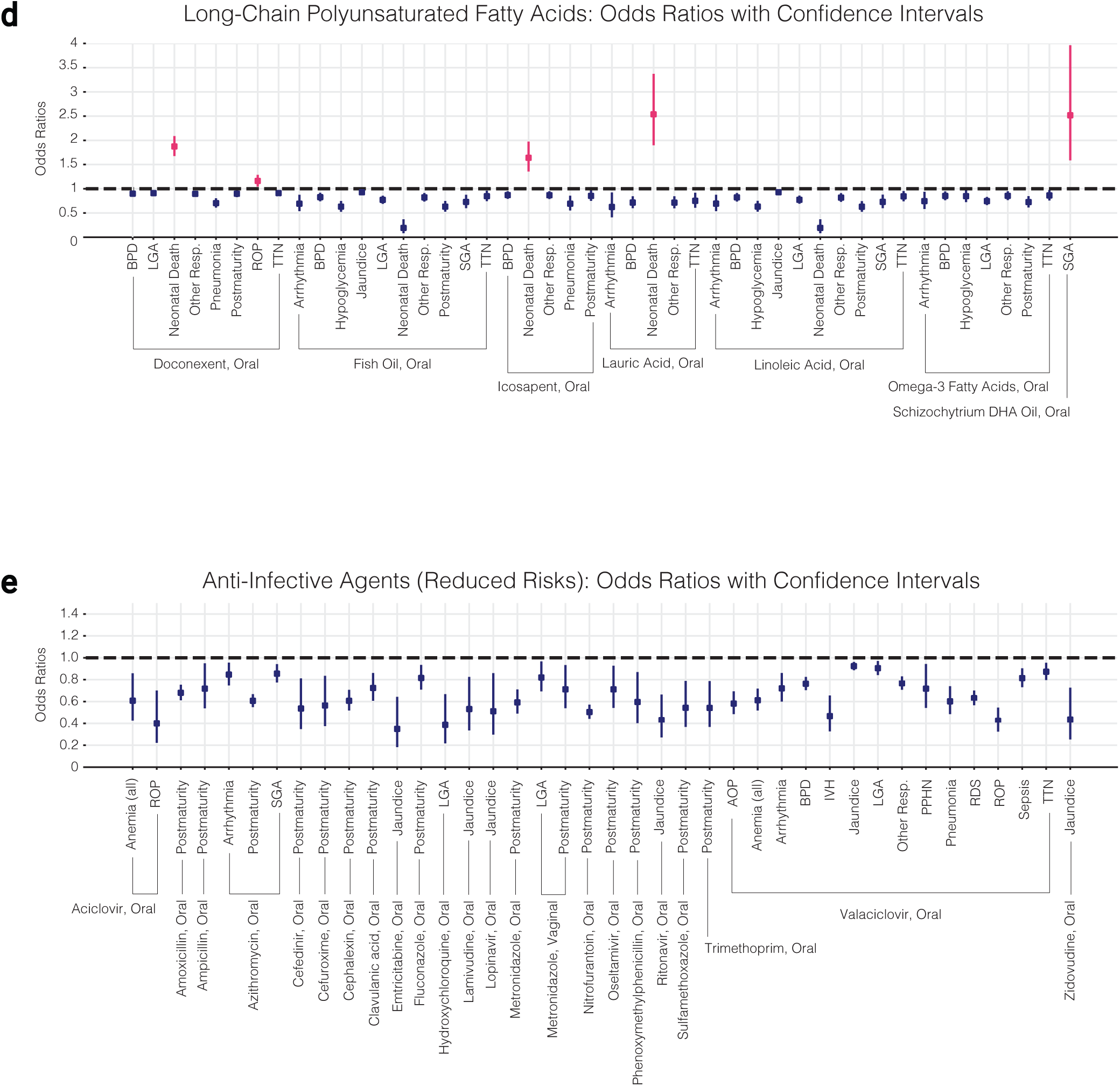
Unadjusted odds ratios for six different medication groups. (a) Antidepressants. (b) Opioids. (c) Antiemetics. (d) Long-Chain PolyUnsaturated Fatty Acids in Vitamins & Combinations Group. (e) reduced risk odds ratios in the Anti-Infective Groups.

**Supplementary Figure 8.**
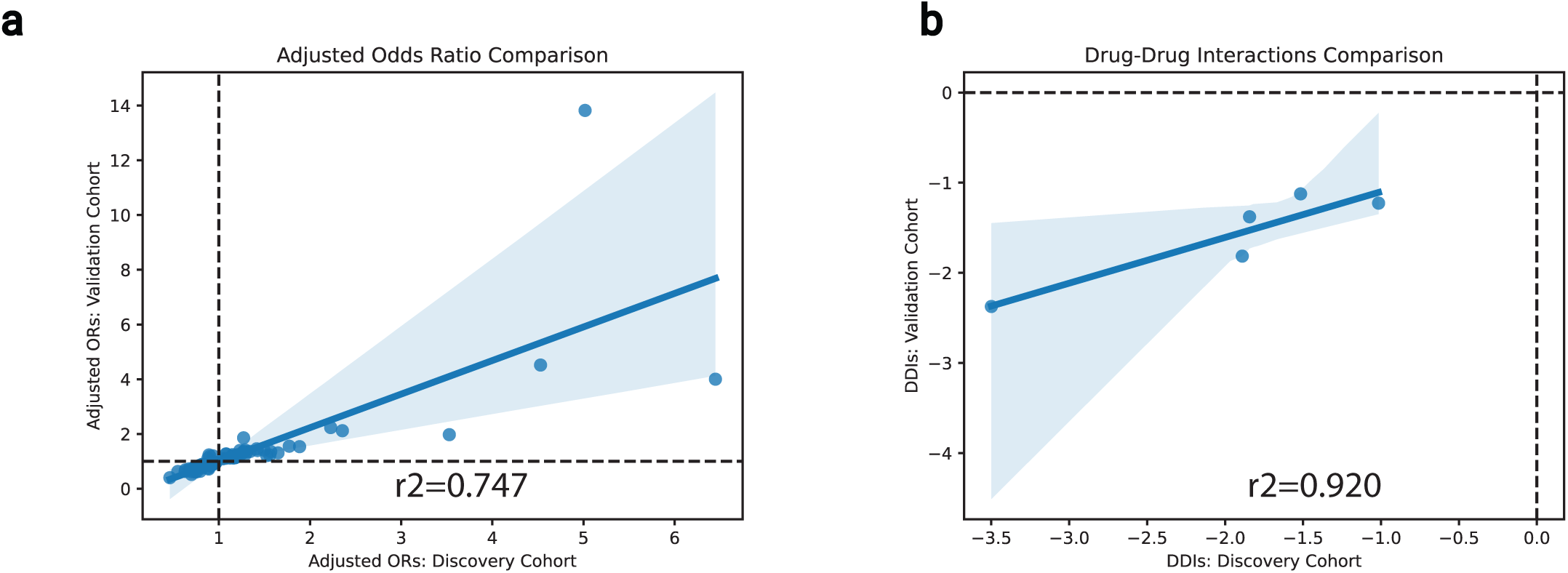
Results from the discovery cohort are correlated with the results from the validation cohort. The results of the association between the two cohorts are matched based on maternal medications and neonatal complications. Scatterplots and Pearson correlation coefficients are generated for these matched results. (a) A scatterplot of significant adjusted odds ratios in the discovery cohort (x-axis) and the validation cohort (y-axis). In total, 81 adjusted odds ratios are matched between the two cohorts, out of 261 from the discovery cohort, and 167 from the validation cohort. The Pearson correlation analysis yields an R2 value of 0.747 with p-value of 1.246e-15. (b) A scatterplot of significant drug-drug interactions (beta3) in the discovery cohort (x-axis) and the validation cohort (y-axis). In total 5 adjusted DDIs are matched between the two cohorts out of 72 from the discovery cohort and 15 from the validation cohort. The Pearson correlation analysis yields an R2 value of 0.920 with p-value 2.682e-2.

**Supplementary Figure 9.**
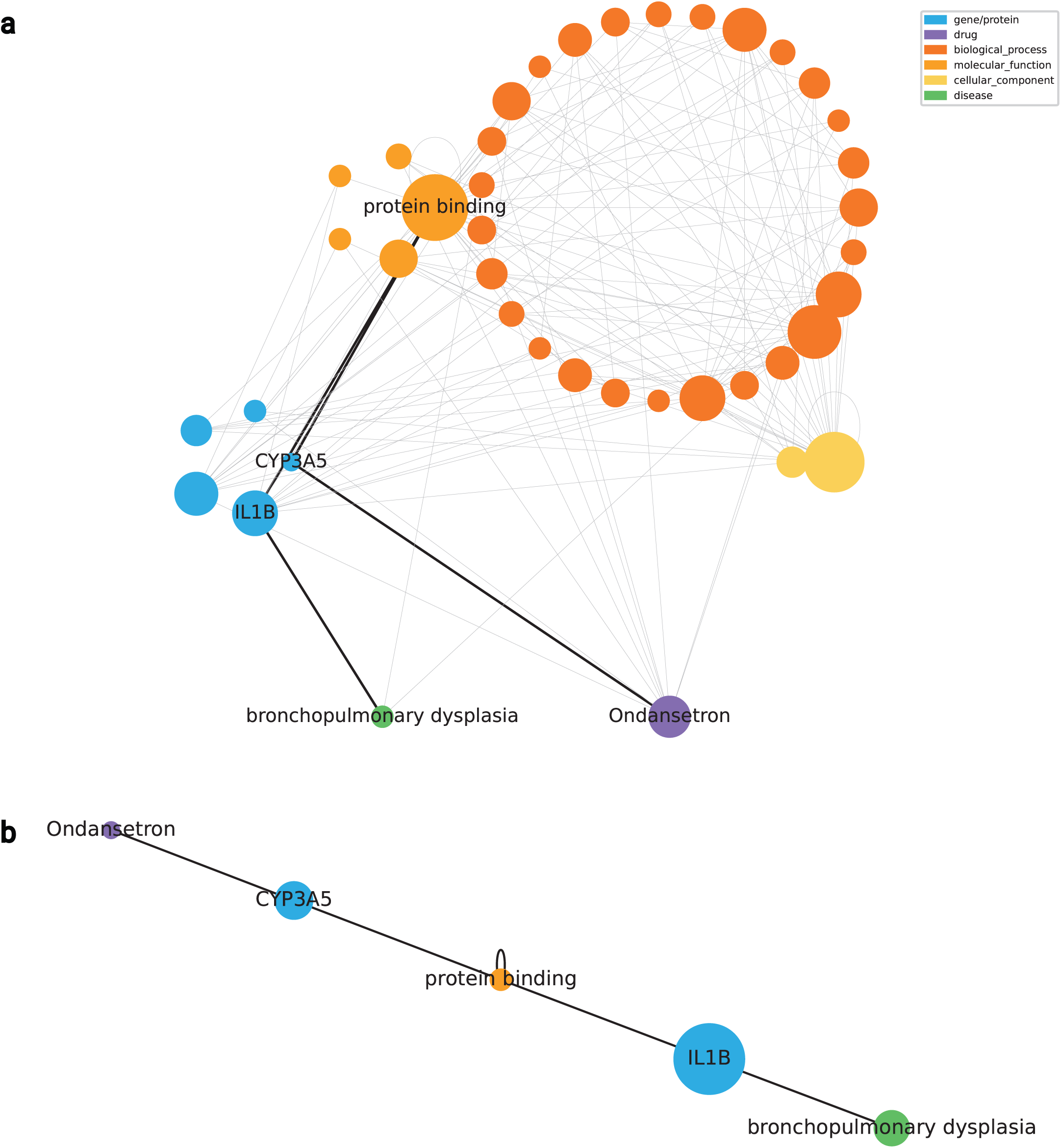
Potential Underlying Mechanisms of Ondansetron Effects on Bronchopulmonary Dysplasia with Inclusion of All Biological Nodes. (a) Hypothesized associations between maternal medications and neonatal complications are generated by combining PregMedNet results with a Knowledge Graph. This is an expanded version of Figure 6. While Figure 6 includes only protein/gene nodes, this subgraph depiction includes all biological node types as intermediate nodes between the disease and the medication of interest. The figure depicts the shortest paths between bronchopulmonary dysplasia and ondansetron. The subgraph contains 60 four-hop paths and 41 nodes, including the medication and disease nodes. (b) The path involving IL-1B and CYP (CYP3A5) is also found in this subgraph, but IL-1B and CYP are connected by a ‘protein binding’ node.

## Notes

### Author Declarations

The Institute of Review Board (IRB) at Stanford University gave ethical approval for this work.

### Summary of Updates

In this updated version, we have: (1) Updated the website URL (https://pregmednet.stanford.edu/), moving the platform from Google Cloud to a Stanford internal server for improved stability and access. (2) Revised the main article to emphasize the utility of claims databases and machine learning approaches for perinatal medication safety research, whereas the earlier version focused more heavily on the results themselves.

